# The impact of digital health insurance for low-income women in Kenya

**DOI:** 10.1101/2023.07.07.23292292

**Authors:** R. de Groot, A. Abajobir, C. Wainaina, E. Sidze, M. Pradhan, W. Janssens

## Abstract

**Objective:** This study evaluates how a subsidized, mobile phone-based health insurance program affected insurance uptake, healthcare utilization and health expenditures for low-income women and their family members in Western Kenya. The program, targeting pregnant women and mothers of children below age four, addressed both demand- and supply-side constraints, providing subsidies through mobile money and support in digital registration while upgrading selected facilities and digitally training community health workers.

**Methods:** The research was based on a cluster-RCT conducted between 2019 and 2021 in 24 villages in Kakamega County. After a baseline survey, 240 households (more than 1,300 individuals) were interviewed every week during 18 months to collect detailed financial and health data while the program was rolled out in the treatment communities, moving to phone-based interviewing after the onset of COVID-19.

**Results:** The intervention had a significant impact on individual insurance uptake of 65.8 percentage points (from a baseline control mean 18.9 percent). We find weak positive impacts on formal healthcare utilization, and substantial increases in financial coverage of medical costs and associated reductions in out-of-pocket expenditures, particularly for medicines. Results are strongest for women, young children and individuals living closest to the clinics. Dynamic analyses show that impacts become increasingly pronounced over time, suggesting that women may need some time to get used to the digital insurance scheme.

**Conclusion:** The program not only reduced the costs of enrolment, but also eliminated other (administrative, logistical, trust) barriers. The introduction of the scheme by trusted local agents, the hands-on assistance with the digital registration procedures at women’s homes, and support in retrieving the necessary documentation such as children’s birth certificates, have likely all contributed to the high enrolment rates, thereby improving access to good-quality care. Digital insurance has the potential to substantially enhance universal health coverage and financial protection for poor households.

## 1. INTRODUCTION

Despite the international commitment to reach Universal Health Coverage (UHC), more than half of the world’s population still lacks access to health care of sufficient quality (World Health Organization & World Bank, 2021) and about 100 million people fall into extreme poverty each year due to ill-health, particularly in low- and middle-income countries (LMICs) (Wagstaff et al., 2018). Many health systems still rely on substantial out-of-pocket payments (OOPs), which restricts access for low-income households. Providing access to affordable health insurance might relieve some of the barriers to UHC, but the uptake of insurance remains low – particularly among poor households (Hooley et al., 2022). Most efforts to increase enrolment so far have produced only modest effects on demand, even when premiums were highly subsidized (Banerjee et al., 2021; Capuno et al., 2016; Thornton et al., 2010; Wagstaff et al., 2016).

One reason for the low uptake may be due to the administrative burden of registration; another reason may be related to quality of health services (Das & Do, 2023). Many insurance schemes do not simultaneously and effectively address the low quality of healthcare (Bonfrer et al., 2018). As a result, benefit packages often provide low value for money, and households may decide not to renew their insurance after having received services of disappointing quality (Duku et al., 2018). Especially low-income households may be deterred by the trade-off between immediate and upfront premium payments versus an uncertain need for health services in the future – especially if the care might be of insufficient quality, and even if their welfare gains from financial protection alone are high (Barnes et al., 2017).

Inequities in access to health care are shaped not only by socioeconomic status but also by gender. Women in Sub-Saharan Africa bear a disproportionate share of the world’s burden of disease, as they account for more than half of the world’s female deaths due to communicable diseases, maternal mortality and nutritional deficiencies (WHO, 2012). Women’s low empowerment is one of the contributing factors. Many African health systems still rely on substantial out-of-pocket payments (OOPs), which restricts access for low-income households. This puts women at a disadvantage compared to men if they are financially dependent on their husbands and need their husband’s consent to seek health care and purchase health services.

This paper evaluates the impact of a digital (mobile-phone based) health insurance program that provided support to enroll in fully subsidized health insurance while simultaneously enhancing the quality of care in selected health facilities. As such, it addressed both demand- and supply-side constraints to insurance uptake. Our research objective was to assess its impact on enrolment, utilization and financial protection using unique high-frequency financial and health data collected on a weekly basis over the period of one and a half year from men and women separately. These data enable us to provide high-detail insights into seasonal patterns of illness incidence, health-seeking behaviors by gender, and out-of-pocket expenditures in relation to fluctuating incomes.

The program targeted low-income women of reproductive age and their family members in Kenya with the aim of enhancing their access to good quality healthcare. It encompassed several digital components. Most notably, it consisted of fully subsidized access to the National Hospital Insurance Fund (NHIF) for the target women as well as their husbands and children. The insurance policy was registered on women’s own sim-cards, thereby increasing their ability to visit a healthcare facility when needed. This was expected to enhance women’s agency and empowerment with respect to their own and their children’s health. To lower administrative and logistical barriers to enrollment, women were approached in their communities, reducing their need to travel to the insurance office, and supported in the administrative procedures.

The mobile-phone based insurance subsidy was embedded within a broader package of health systems changes, including quality improvement support for selected healthcare providers; a digital training package for local community health workers (CHWs); a digital household registration survey tool to aid CHWs during their community visits; and a digital health platform (called M-TIBA) that connected patients, healthcare providers and the NHIF on a real-time basis to stimulate data sharing and enhance efficiency, transparency and accountability in the system. CHWs were closely involved during the enrolment of women on the insurance scheme to raise awareness on NHIF and encourage the women to sign up. This was expected to further enhance uptake, due to greater trust. The impact evaluation focuses only on the effects of the subsidized, digital health insurance coverage combined with the enrolment support, because households in both treated and control communities could benefit from the quality upgrades in the selected clinics and the improved training of CHWs in the study area.

The experimental design of the impact evaluation was based on a matched-pair, longitudinal, cluster randomized controlled trial (RCT), with randomization at the village-level. The study sample includes 24 villages, 240 households, and more than 1,300 individuals. After a baseline survey at the end of 2019, all adults in the households – both women and men – were interviewed individually and separately on a weekly basis to collect detailed diaries data on all the health events that occurred in the household in the past week (including symptoms, consultations, provider choice, treatments and health expenses of adults as well as children) and all their financial transactions (including incomes and expenditures, loans and savings, and gifts and remittances). From June 2020 onwards, the subsidized insurance scheme was rolled out in the treatment communities. An endline survey in June 2021 completed the data collection.

We aggregated the individual weekly data to the month-level and estimated the intent-to-treat effects (ITT) and average treatment effects on the treated (ATT) using ANCOVA specifications, and instrumental variable techniques using village treatment assignment as instrument. We test for heterogeneous effects by gender, age, socioeconomic status, and distance to the healthcare facility.

Our key findings show that the program was successful in increasing enrolment in NHIF: the share of individuals in the treatment households who were enrolled in the insurance scheme increased with 65.8 percentage points from a baseline of 18.9 percent. However, this success did not come naturally. A major hurdle to enrolment turned out to be the NHIF requirement of birth certificates for children and national ID-cards for adults. Many households in the low-income communities did not have a birth certificate for each of their children and program staff invested heavily to relieve this constraint. The program could not support target women without an ID-card, however, and they were dropped from the study sample. The conclusion delves further into the implications thereof both for policy-making and external validity.

We find a significant impact of the subsidized health insurance scheme on the probability of seeking care when ill or injured, with on average 13.8 percentage points up from a baseline mean in the control group of 49.3 percent. This impact is driven by visits to formal, program-selected providers, and strongest for adult women – even when focusing on healthcare seeking for reasons other than reproductive, maternal, neonatal and child health (RMNCH), and taking into account that at baseline they were already twice as likely to seek formal care compared to adult men.

Impacts on healthcare utilization are also much larger for households in treatment villages linked to the private rather than the public program-selected health facility. This is – at least in part – due to the greater distances that households had to travel to public hospital. Indeed, we find that the impact on seeking formal care is strongly dependent on the distance to the program-selected healthcare provider.

In line with the high enrolment rates, the digital insurance scheme significantly enhanced financial coverage of medical costs for illness and injury, with an increase of 30.9 percentage points from a baseline of 8.3 percent among control households. The increases in financial coverage were most pronounced for women and girls. Out-of-pocket health expenditures for illness and injury decreased concomitantly, with on average KES 797 annually for the insured, or KES 231 per insured formal consult. The likelihood of incurring catastrophic health expenditures (CHE) decreased concomitantly with 7.4 percentage points from a baseline control mean of 10.1 percent.

Our research builds on three strands in the literature. First, it adds to the literature that examines the impact of health insurance in LMICs on health care utilization, and out-of-pocket expenditures (OOPs), where we specifically focus on the gender dimension. Systematic reviews generally find substantial impacts on healthcare utilization, as well as positive effects on out-of-pocket expenditures although findings are not always conclusive and dependent on the context (Das & Do, 2023; Acharya et al., 2013; Spaan et al., 2012). In line with those studies, we find a substantial positive effect on utilization, OOPs and financial protection. Importantly, we show that these effects are most pronounced for women and girls – even though men and boys are just as likely to become enrolled in the digital health insurance scheme.

Second, it contributes to our understanding of how to increase the uptake of health insurance in LMICs. Previous studies have found limited impacts of subsidizing insurance or providing information on enrolment in Vietnam and the Philippines (Capuno et al., 2016; Wagstaff et al., 2016). A study in Ghana found that full subsidies increased enrolment in the national health insurance scheme significantly with 54 percentage points from a baseline of about 20 percent (Asuming et al., 2018). In Nicaragua, on the other hand, subsidizing insurance for informal sector workers only attracted 20 percent new enrolments, and the savings in OOPs through insurance were not enough to recover the full insurance premium for most households (Thornton et al., 2010). Similarly, in a paper closely related to ours, Banerjee et al. (2021) study different treatments to increase insurance coverage in Indonesia. They find that subsidies and registration assistance increased enrolment, but not an information treatment. In one of their treatments, a full (100%) subsidy increased enrolment by less than 20 percentage points, far from universal coverage. These studies shows that there are substantial non-price barriers to enrolment and health insurance. Our study shows that the offer of (fully subsidized) digital insurance on women’s own sim-cards in combination with hands-on assistance to navigate the administrative requirements and registration procedures, and carried out by trusted agents within the community, can raise individual enrolment levels up to 85 percent.

Third, our paper fits within the emerging literature on the potential of digital technology and mobile money (fintech) to advance financial inclusion. Mobile technology is regarded as one of the key drivers of financial inclusion in recent years (Demirgüç-Kunt, Klapper, et al., 2017) and Kenya has been at the forefront of the mobile money revolution (Suri & Jack, 2016). An increasing literature shows how digital financial inclusion, including access to digital savings and lending, can support women’s economic empowerment. In addition, MPESA has been shown to significantly enhance households’ ability to cope with health and other shocks through informal risk-sharing between households (Jack & Suri, 2014). Nevertheless, there is room for improvement in risk management through formal insurance schemes, as the most vulnerable individuals – including women and the poor– may be excluded from effective social support networks (Geng et al., 2018). The digital intervention offered by the program bridges the gap between digital finance and insurance subsidies targeting low-income women.

The main methodological contribution of our study is our ability to explore at a granular level the health care decisions of households. Using our weekly health diaries, covering a period of 18 months, we are able to assess the impact of subsidizing insurance on health-seeking behavior at a variety of providers, as well as OOPs and catastrophic spending on a high-frequency basis. Das et al. (2012) clearly show how health surveys are hampered by a recall bias. The longer the recall period, the more likely respondents are to remember only severe illnesses and injuries, or health events that required high health expenditures. However, Nelissen et al. (2020) show that minor but regular health events might represent substantial portions of annual OOPs; standard surveys may miss out on as much as half of OOPs or more. The set-up of our weekly diaries ensures that all those events are accounted for. Note that Banerjee et al. (2021) also use high-frequency data, but their data only covers the supply side, as insurance claims are only observed if individuals chose to seek health care for a health problem. In our case, we observe the full universe of health problems in a sample population with the corresponding decisions of whether people sought health care, where, which provider, how much they paid and whether their visit was covered by insurance. This depth of information allows us to identify if and how health insurance can make a difference in low-income populations. For example, we show that impacts of the program on utilization steadily increases over time, and that the effects on OOPs are driven mostly by a reduction in spending on medication.

## 2. CONTEXT AND INTERVENTION

### 2.1 Study setting

Although being classified as a middle-income country in 2014, Kenya remains among the 25% poorest countries in the world, affected by social and health inequalities. More than one-third of Kenyans had an income below the poverty line (1.9 USD/day) in 2015 (World Bank, 2021). Inequalities in access to healthcare, in particular maternal and child health care, are still rampant, despite major improvements made through targeted policies over the past few years (United Nations Department of Economic and Social Affairs, 2015). For instance, according to the 2014 Kenya Demographic and Health Survey (KDHS), the maternal mortality ratio has marginally reduced to 362 per 100,000 live births, not statistically different from the figures reported in 2008-2009 (Kenya National Bureau of Statistics et al., 2015). While a substantial under-five mortality reduction was achieved with a drop from 115 per 1000 live births in 2003 to 52 per 1000 live births in 2014, it is still two-folds higher than the SDG target.

The impact study was carried out in Khwisero sub-county of Kakamega County. The reported under-5 mortality ratio in Kakamega is 64, slightly above the national average. The most recent figures put Khwisero’s population at 113,000 people with a very rural character, with nearly 85% of households in the sub-county engaged in agriculture (Kenya National Bureau of Statistics, 2019a, 2019b).

Enrolment in Kenya’s National Health Insurance Fund (NHIF) stood at 14% in 2017, mainly covering civil servants and other formal sector workers as well as a limited number of low-income households through the health insurance subsidy for the poor (HISP) program, targeting orphans and other vulnerable children. Most informal sector workers choose not to enroll in the voluntary insurance scheme of NHIF with monthly household premiums of 500 KES (Barasa et al. 2018). To further enhance UHC, the Government of Kenya recently included it as one of its ‘Big Four Agenda’-action points (Wangia & Kandie, 2019) with the objective to achieve a 100% cost subsidy for essential health services and to reduce out-of-pocket health expenditures by half. As part of these efforts, Linda Mama was implemented through the NHIF starting in 2017, providing a basic package of free ante-natal care, skilled delivery services, neo-natal care, and post-natal care in all NHIF-empaneled healthcare providers. Low-cost health insurance schemes, including eHealth and mobile health (mHealth) services, are among the other strategies piloted to achieve this goal.

### 2.2 The intervention

The Innovative Partnership for Universal Sustainable Healthcare (i-PUSH) program has been implemented by the non-governmental organizations Amref Health Africa and PharmAccess Foundation since 2017 to support the Kenyan government in its efforts towards reaching UHC. The i-PUSH program aims to empower low-income women of reproductive age and their families by enhancing their access to healthcare through innovative digital tools. This section describes the intervention in more detail.

The first component, which is the focus of our evaluation, was the subsidized digital health insurance scheme^1^. The program provided free access to the SupaCover scheme of the NHIF, which includes a comprehensive package of out- and in-patient healthcare services at NHIF- empaneled public and private providers. As such, it complemented the basic package of free services for reproductive, maternal, neonatal and child health (RMNCH) care at public providers through the Linda Mama scheme. The digital insurance component was implemented through the M-TIBA digital health platform that connects patients to providers. Program staff first identified households belonging to the target population with the aid of CHWs, who have good knowledge of their assigned catchment area of approx. 100 households. Households were eligible if they were living in a low-income community and included at least one woman of reproductive age (15-49 years old) who was either pregnant or with a child below age four. Women in the identified households were subsequently visited by an M-TIBA agent, accompanied by the respective CHW to facilitate introductions. Agents and CHWs would give information about the digital insurance to each woman, i.e. the offer of a full year of fully subsidized NHIF cover (SupaCover) for her and her family members registered through a mobile phone on her sim-card. If a woman was interested – and conditional on her having her own sim-card – the agent would assist in the digital registration procedure. If she needed more time to consider the offer, to get a sim-card, or to collect the necessary documents, a second visit was planned. As an NHIF requirement, an ID-card or birth certificate was necessary to register adult and child household members, respectively. Since a considerable number of households did not have a birth certificate for their child, the program staff also assisted in obtaining such certificates from the County. Once NHIF had approved the digital registration, coverage was activated to start at the first day of the next month with a duration of one year. Many participants also desired to have a physical NHIF card, which was later provided upon request.

The second component was targeted towards CHWs. The program enhanced their training using a mobile phone-based training tool, which focused on raising awareness about health insurance and on improving RMNCH knowledge, providing information e.g. on the importance of timely antenatal care, danger signs during pregnancy, nutrition requirements of children under 5, and family planning. The improved CHW training in turn was expected to also improve health knowledge and behavior of the target women through their contacts with the CHWs. CHWs also conducted a detailed household mapping using a specialized data collection tool, on the basis of which the target beneficiaries for the digital insurance were identified.

The third component focused on selected healthcare providers. The program enhanced the quality of care in selected healthcare providers based on the SafeCare approach, an IEEA accredited standards-based stepwise quality improvement methodology (Johnson et al., 2016). SafeCare consists of a professional quality assessment after which the clinics are encouraged to develop a quality improvement plan (QIP). It does not provide resources to the healthcare providers directly, but it provides technical assistance in the development and implementation of the QIP. The healthcare providers were also enrolled onto the M-TIBA platform to facilitate the collection of health utilization data.

Women enrolling on NHIF through the digital program were requested to choose one healthcare provider as their preferred facility from the list of NHIF-empaneled providers. Program agents encouraged women to select as their preferred provider one of the facilities that had participated in the SafeCare quality improvement program, but this was not compulsory.^2^ Upon arrival at the NHIF healthcare facilites, program enrollees would use their mobile phones and show their ID-card to be registered in the digital M-TIBA system, or they could present the physical NHIF card.

## 3. RESEARCH METHODOLOGY

### 3.1 Experimental design

Our experimental design is based on a cluster-randomized control trial approach. We first randomly selected 24 villages in the catchment area of four healthcare providers (six per provider). We then matched villages in 12 pairs, based on village-level characteristics and aggregated household characteristics from a baseline survey conducted at the end of 2019, and randomly assigned one village per pair to the intervention and one to the comparison group. In each village, 10 households were randomly selected from the target population to reach a baseline sample size of 240 households (120 treatment households and 120 control households), encompassing more than 1,300 individuals.

More precisely, at the start of the study in September 2019, four healthcare providers in Khwisero sub-county had attained NHIF level 4, and were hence eligible for NHIF-empanelment, providing both out- and in-patient care. Of these, three were private and one was public^3^. They all served low-income communities, and were invited to participate in the SafeCare quality improvement process, funded by the program. Six villages located in the catchment areas of each of these four facilities were randomly selected from a list of all villages in their catchment areas.

In each village, the program manager together with the local CHWs provided a complete listing of households. Based on household demographics and pregnancy information, eligible households were identified in line with the program eligibility criteria. Households eligible for the study included those with at least one woman of reproductive age (aged 18-49)^4^, who: a) had at least one child below 4 years living with her at baseline; or b) was pregnant at baseline. Ten eligible households were randomly selected from this list in each village to be included in the study sample. Additional eligible households per village were sampled as replacement for refusals and dropouts.

Initially, the study sought a 50-50 allocation between households with a pregnant woman and households with a child under 4 years old. After the household listing exercise, it became clear that there were too few pregnant women in each village to fulfill this criterion. Therefore, all pregnant women were included in the study sample up to five per village, and we randomly sampled additional households with children under 4 years old until the cluster size (10 households per village) was achieved. The selected healthcare providers and location of the sampled household is provided in Figure 1.

**Figure 1.**
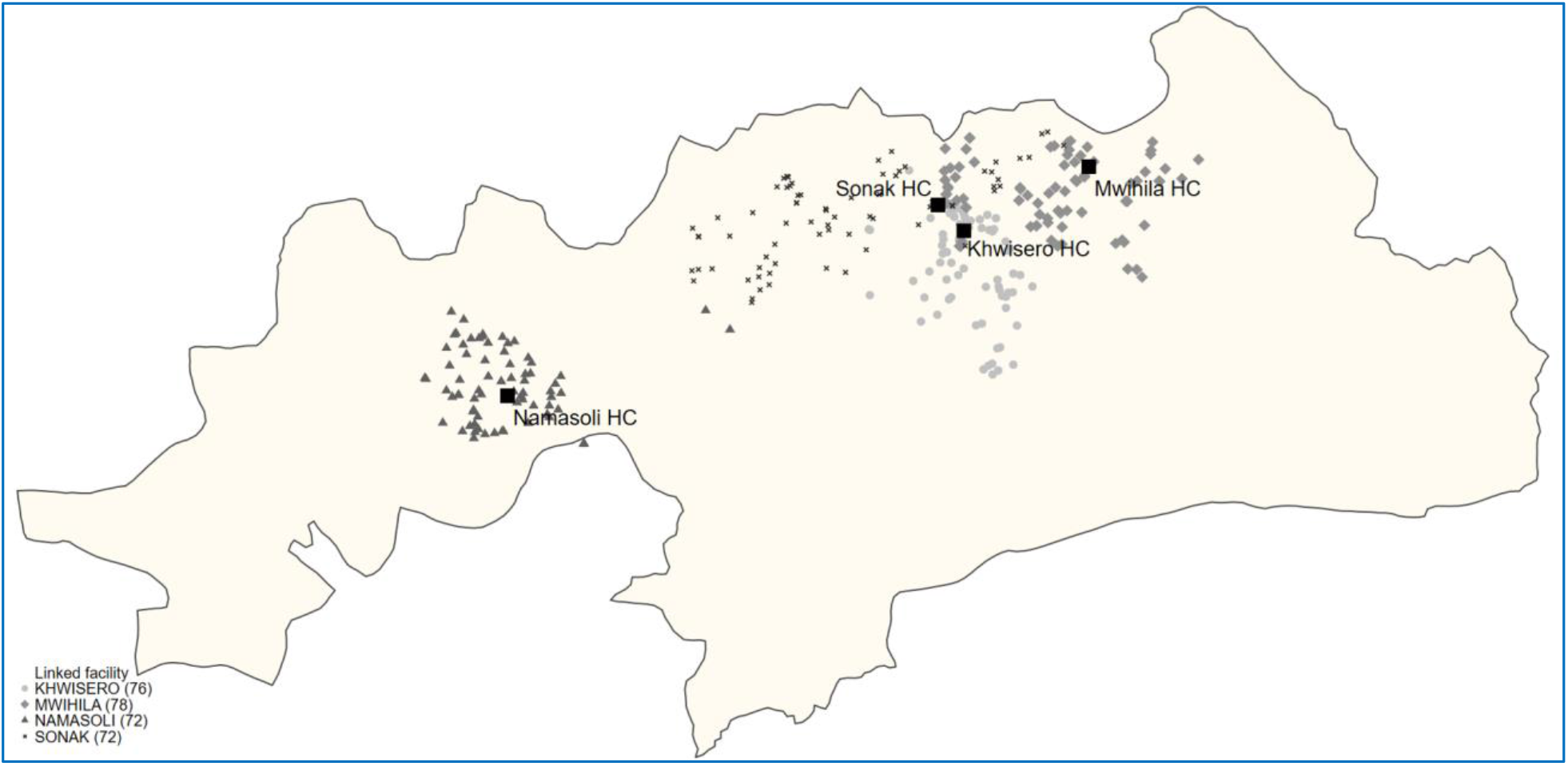
Map of the study site (with GPS locations of study households and originally selected healthcare providers) Note: Private Sonak Health Center (HC) and private MWhila HC were initially invited but ultimately not included in the i-PUSH intervention. Private Namasoli HC and public Khwisero HC were included.

Next, we conducted a baseline survey with the sampled households and a village survey with key informants to obtain information for the village-level randomization matching procedure. We matched villages in 12 pairs that were as similar as possible to each other (Imai et al., 2009).^5^ Our matching indicators included village-level indicators (demographics, infrastructure, availability of health services) and household-level indicators, aggregated at the village-level (female educational attainment, share of women earning income, membership of savings groups for women, mobile phone ownership for women, wealth, health care utilization, health expenditures, health insurance).

Randomization was blocked at the health facility level to ensure that each health facility served an equal number of treatment and control villages. Thus, each village was matched with one of the other five villages in the catchment area of its nearest health facility.

Assignment to the treatment or the control group was carried out by the research team during a public ceremony in the presence of key stakeholders, local liaison persons and village representatives on March 12^th^, 2020 – a few days before the onset of the COVID-19 pandemic in Kenya. Consent for the procedures was obtained from local government officials before the random assignment. During the randomization ceremony, papers with paired village names were folded and put in a bag; two village representatives from each paired village discussed whom would pick the paper; and after the other group members verified that the names could not be seen, one paper was picked, and a coin was flipped to decide which group the picked village belonged to. The process of choosing the folded paper and flipping of the coin was repeated for all paired villages.

The roll-out of the digital insurance component, initially scheduled to start shortly after the public randomization ceremony, was delayed until June 2020 due to the stringent COVID-19 lockdown measures implemented from March 15^th^ onwards. The 12 treatment villages were visited sequentially, starting in June. Enrolment of individual households in treatment villages was completed by the end of September 2020.

Contrary to original plans, only two of the four selected healthcare providers enrolled in the SafeCare quality improvement program and onto the M-TIBA digital health platform. One private facility was not included because of organizational challenges; another private facility was dropped because it was located 500 meters away from the public facility, with overlapping catchment areas (see Figure 1). Households in the treatment communities were therefore encouraged to choose one of the two remaining program-selected healthcare providers as their preferred NHIF provider. We investigate the effect of geographical accessibility on utilization of the scheme in our heterogeneity analyses.

### 3.2 Data collection

The baseline survey in October-November 20218 collected information on household demographics, socio-economic indicators, food consumption indicators, financial inclusion, participation in community networks, as well as self-assessed health status, health-related knowledge and behavior, health care utilization and health expenditures, maternal health, mental health, intra-household decision-making processes and gender dynamics. In December 2020, we conducted a midline household survey, similar to the baseline survey, and in June 2021 we concluded the data collection with an endline survey (see Figure 2).

**Figure 2.**
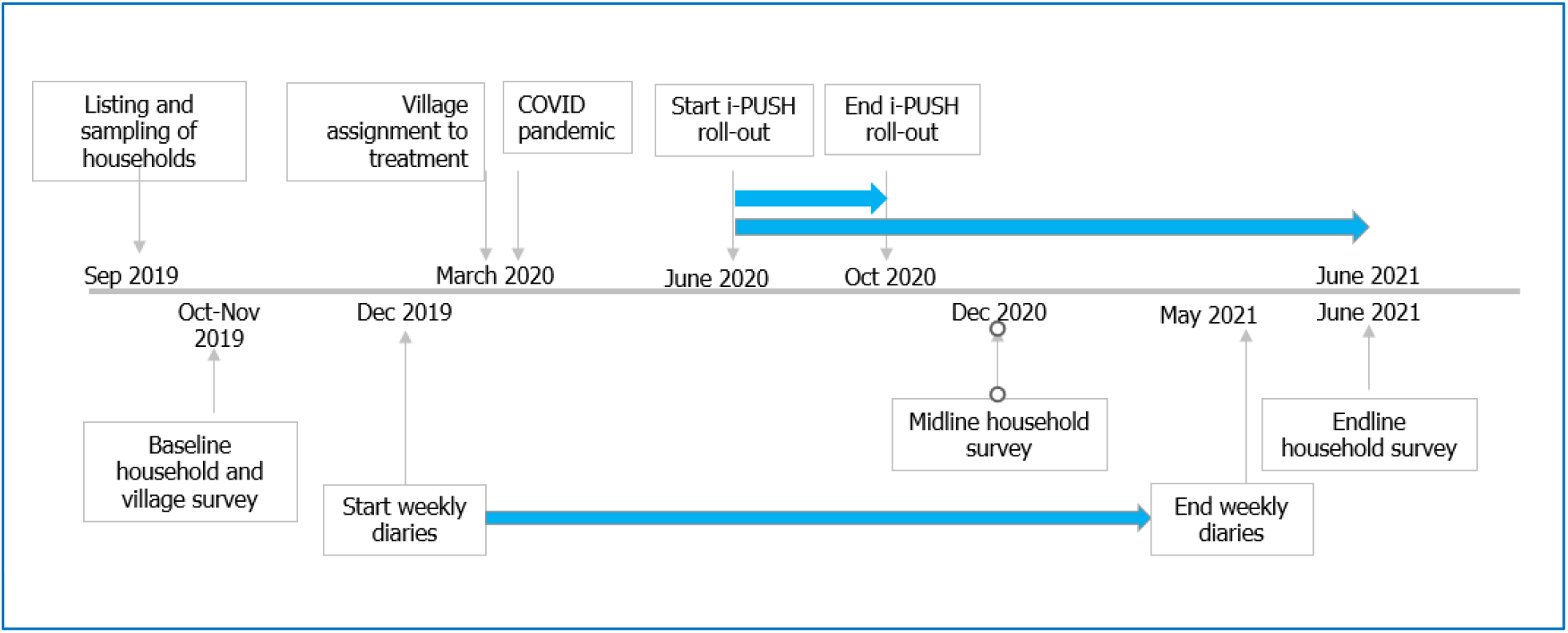
Timeline of data collection and intervention.

Two weeks after the baseline data collection was completed, we started collecting weekly financial and health diaries and continued the diaries interviews for 18 months, from December 2019 until the end of May 2021. The financial diaries recorded all financial transactions from the seven days prior to each interview, including income and expenditures, loans, gifts and remittances given and received, and savings. Respondents to the financial diaries encompassed all economically active adult household members able to participate in the interviews (e.g. excluding the very old-aged or disabled). Men and women were interviewed separately and in private about their own financial transactions.

The health diaries provided a detailed picture of the prevalence of illnesses and injuries, as well as preventive and curative healthcare utilization. Through the health diaries, we collected data on all health events that occurred to any of the household members in the seven days prior to each interview. Respondents were the adults from the financial diaries who responded for themselves as well as for adult household members who were absent, old-aged or disabled; while an informed adult, usually the mother, would respond for the children in the household. Collected health data included symptoms, whether any health care was sought, which health provider was visited, which health services were received, out-of-pocket health expenditures, date of onset of the symptoms and date of provider visit(s).

After the first case of COVID-19 was registered in Kenya in March 2020, the government implemented stringent movement restrictions. Field work switched from in-person interviewing to phone interviews. Due to the strong built-up rapport and trust between the field workers and respondents during the three months preceding the switch, this transition occurred with minimal disruptions (Janssens et al., 2021).

### 3.3 Study sample and response rates

The study enrolled 240 households during the baseline survey. After the baseline survey, but before the roll-out of the intervention, it became clear that some target women in the treatment group (N=11) did not have a valid national ID-card, which was a compulsory condition to obtain NHIF coverage. These women and their household were therefore replaced in the study sample with households from the replacement list.^6^ In addition, during the seven months between the baseline survey and the roll-out of the intervention from June onwards, 40 households (27 in the treatment and 13 in the control group) had dropped out due to other reasons, mostly because of COVID-19-induced relocation. The treatment and control samples were therefore replenished in August 2020 using the remaining list of replacement households, and the newly included households participated in a short version of the baseline survey.

After replenishment, the total study sample included 243 households (123 treatment households and 120 control households). Households that dropped out of the sample before August 2020 are excluded from the analysis. By the time of the endline survey one year later, 232 households (118 in the treatment group and 114 in the control group) were retained in the sample. Attrition rates were hence low at 3.3 percent (1.7 percent and 5.0 percent in the treatment and control group, respectively). These 232 households (with 1,276 individual household members) constitute our balanced main analysis sample, for which we have collected baseline, midline and endline data, as well as weekly diaries data. See Appendix A Figure A-1 for a detailed description of the sampling methodology and sample size.

We performed several balance checks to verify comparability of the treatment and the control group, given the changes in the study sample over time. We find very few statistically significant differences between the characteristics of the treatment and the control group in our final analysis sample (Table 1). Likewise, baseline differences between households in the analysis sample and those dropping out before August 2020 were minimal. Given the limited imbalance, we are confident that the randomization was successful, and that the replenishment did not lead to a selective sample.

**Table 1.** Descriptive statistics at baseline (by treatment status)

| Variables | Total |  | Control |  | Treatment |  | p-value of the difference |
| --- | --- | --- | --- | --- | --- | --- | --- |
|  | Mean | N | Mean | N1 | Mean | N2 |  |
| **Panel a. Demographic characteristics** |  |  |  |  |  |  |  |
| # of household members | 4.879 | 232 | 4.965 | 114 | 4.797 | 118 | 0.533 |
| # of members aged 0-5 years | 1.302 | 232 | 1.316 | 114 | 1.288 | 118 | 0.793 |
| # of members aged 6-12 years | 1.034 | 232 | 0.991 | 114 | 1.076 | 118 | 0.549 |
| # of members aged 13-18 years | 0.556 | 232 | 0.570 | 114 | 0.542 | 118 | 0.825 |
| # of members aged 0-18 years | 2.892 | 232 | 2.877 | 114 | 2.907 | 118 | 0.907 |
| # of members aged 19-64 years | 1.957 | 232 | 2.035 | 114 | 1.881 | 118 | 0.104 |
| # of members aged 65 and over | 0.030 | 232 | 0.053 | 114 | 0.008 | 118 | 0.092* |
| Head: age in years | 37.078 | 232 | 38.088 | 114 | 36.102 | 118 | 0.233 |
| Head: 1 if female | 0.250 | 232 | 0.237 | 114 | 0.263 | 118 | 0.627 |
| Head: married | 0.914 | 232 | 0.921 | 114 | 0.907 | 118 | 0.680 |
| **Panel b. Socio-economic characteristics** |  |  |  |  |  |  |  |
| Head: Completed primary education or higher | 0.638 | 232 | 0.623 | 114 | 0.653 | 118 | 0.600 |
| Any hh member engaged in own business | 0.220 | 232 | 0.237 | 114 | 0.203 | 118 | 0.601 |
| Any hh member in engaged in own farm work | 0.091 | 232 | 0.114 | 114 | 0.068 | 118 | 0.252 |
| Any hh member engaged in casual labor | 0.504 | 232 | 0.509 | 114 | 0.500 | 118 | 0.892 |
| Any hh member engaged in wage work | 0.211 | 232 | 0.158 | 114 | 0.263 | 118 | 0.096* |
| Weekly hh income from work (KES) | 3,600 | 232 | 4,701 | 114 | 2,536 | 118 | 0.008*** |
| Average wealth index | -1.007 | 232 | -1.248 | 114 | -0.774 | 118 | 0.306 |
| Share of hh in low wealth tercile | 0.466 | 232 | 0.482 | 114 | 0.449 | 118 | 0.695 |
| Share of hh in middle wealth tercile | 0.384 | 232 | 0.421 | 114 | 0.347 | 118 | 0.171 |
| Share of hh in high wealth | 0.151 | 232 | 0.096 | 114 | 0.203 | 118 | 0.185 |
| Household owns any livestock | 0.763 | 232 | 0.798 | 114 | 0.729 | 118 | 0.383 |
| Household owns any cattle | 0.371 | 232 | 0.377 | 114 | 0.364 | 118 | 0.864 |
| Household owns any land | 0.728 | 232 | 0.746 | 114 | 0.712 | 118 | 0.762 |
| Household has agricultural stock | 0.552 | 232 | 0.553 | 114 | 0.551 | 118 | 0.985 |
| Savings in saving group/cooperative | 9,142 | 193 | 5,533 | 107 | 13,633 | 86 | 0.156 |
| Savings at other locations (KES) | 3,134 | 193 | 4,748 | 107 | 1,127 | 86 | 0.063* |
| Total amount of savings (KES) | 12,276 | 193 | 10,280 | 107 | 14,760 | 86 | 0.429 |
| Household has formal loan | 0.223 | 193 | 0.234 | 107 | 0.209 | 86 | 0.693 |
| Household has informal loan | 0.399 | 193 | 0.364 | 107 | 0.442 | 86 | 0.308 |
| Loan amount outstanding (KES) | 3,885 | 193 | 3,411 | 107 | 4,474 | 86 | 0.332 |
| Amount of money lent out (KES) | 1,892 | 193 | 1,521 | 107 | 2,354 | 86 | 0.403 |
| **Panel c. Health-seeking behavior and out-of-pocket health expenditures (OOPs) in the past 12 months** |  |  |  |  |  |  |  |
| Number of times inpatient care | 2.456 | 193 | 2.299 | 107 | 2.651 | 86 | 0.638 |
| Number of times outpatient care | 15.332 | 193 | 16.206 | 107 | 14.244 | 86 | 0.206 |
| Number of times informal vendor | 14.466 | 193 | 14.486 | 107 | 14.442 | 86 | 0.988 |
| Number of times traditional healer | 2.756 | 193 | 2.897 | 107 | 2.581 | 86 | 0.773 |
| Number of times foregone care | 3.570 | 193 | 4.299 | 107 | 2.663 | 86 | 0.069* |
| Total health expenditures (KES) | 8,133 | 232 | 9,733 | 114 | 6,588 | 118 | 0.239 |

Notes: The table reports the mean values for the control and treatment groups for each variable listed in the table, as measured in the baseline survey. The p-value comes from a Wald test of the equality of means with standard errors clustered at the village level. Statistics are based on the analysis sample of households in the balanced panel.

### 3.4 Ethics

All respondents were asked for informed consent before participating in the baseline survey as well as at the start of the diaries data collection. Ethical approval was granted by the Amref Health Africa Ethics and Scientific Review Committee (P679-2019, 8^th^ August 2019), with an amendment for the COVID-19-induced switch to telephone interviews (granted on 21^st^ of April 2020).

### 3.5 Measurement of the outcome variables

The primary outcome variables are related to the enrolment in health insurance, the utilization of health care, and financial protection.^7^ Enrolment in NHIF health insurance was recorded at the individual level in the baseline, midline and endline surveys (recording whether currently insured, and if so, enrolment date and end date of the insurance), and transformed into a monthly binary indicator for NHIF insurance status. We cannot explicitly distinguish between treatment households who enrolled in NHIF themselves or through i-PUSH. We also measure whether the individual was enrolled in any insurance scheme other than NHIF in a particular month. We analyze monthly household level enrolment through an indicator equal to one if any household member was enrolled in NHIF in that month.

Healthcare utilization is recorded at the individual level on a weekly basis in the health diaries.^8^ First, we examine whether an individual had a consultation for any reason at any healthcare provider during an interview week. This could be a consultation for preventive care (such as health check-ups or vaccinations), for curative care (i.e. for treatment of an illness or injury) or for RMNCH care (such as family planning or antenatal care), at any type of healthcare provider. Next, we analyze healthcare utilization at the various provider types separately, i.e. any informal provider (such as a traditional healer or informal drug vendor); any formal provider (such as a doctor, nurse, midwife or pharmacist in their private practice, a public or private clinic/health center/hospital, or a pharmacy); and one of the two program-selected providers. We also analyze healthcare utilization *conditional* on being ill or injured, restricting the dataset to weeks in which individuals reported a health shock.

Financial protection from health risk is measured using three sets of indicators. First, we investigate insurance coverage of consultations at a formal provider, restricting the sample to individual-weeks with a formal consultation, and we examine whether the costs of that consultation were not covered by insurance, (partially) covered by NHIF, fully covered by NHIF. Next, we analyze OOPs, recorded at the individual level in the weekly health diaries, measured in Kenyan Shillings. Our third measure of financial protection is measured as the probability of catastrophic health expenditures (CHE). Following Xu et al. (2014), we measure CHE as OOPs exceeding a household’s capacity to pay (CTP). We calculate OOPs as weekly averages, aggregated at the household-month level. CTP is calculated as a household’s monthly non-subsistence spending (the household’s total expenditures minus the poverty line); this captures the money left for the household to spend after taking care of basic necessities such as food consumption. The CHE variable is a binary indicator, equal to 1 if OOPs are larger than 40% of CTP (with 30%, 20%, and 10% included as robustness checks).

### 3.6 Empirical methodology

We start with descriptive analyses of the dynamics over time, comparing mean outcomes of households in the treatment and control villages over the study period from February 2020 until May 2021.^9^ Our unit of observation in these analyses is the individual-week (except for insurance status that is measured at monthly intervals), which we show as monthly averages in line with the subsequent impact analyses. While these analyses provide visual insights in the dynamics of the outcome variables over time, they do not yield a singular estimate of the impact.

Our main impact specification is based on an ANCOVA model, taking the individual-month as the unit of analysis. We estimate the following equation:

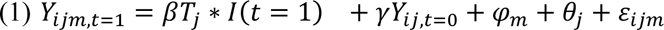

Where . 𝑡 = 0 denotes the pre-intervention period (before program roll-out in June), 𝑡 = 1 denotes the post-intervention period (after program roll-out has been completed, from October onwards)^10^, and 𝐼 is in an indicator function which equals 1 for post-intervention months. 𝑌_𝑖𝑗𝑚,𝑡=1_is the outcome 𝑌 for individual 𝑖 in village *j* in month *m,* measured in the post-intervention period. To this end, we averaged the values observed in the weekly diaries for each month, essentially imputing missing weeks with their month’s average. 𝑌_𝑖𝑗,𝑡=0_ is the average weekly value observed in the pre-intervention period. 𝑇_𝑗_ = 1 if village *j* was assigned to the treatment group, and zero otherwise. 𝜑_𝑚_are month fixed effects and 𝜃_𝑗_ are matched-pair fixed effects (our randomization indicators). Standard errors are clustered at the village-pair level (de Chaisemartin & Ramirez-Cuellar, 2020). The coefficient of interest is *β* which represents the intent-to-treat (ITT) impact of the digital insurance program on outcome 𝑌_𝑖𝑗_, which estimates the impact of living in a village assigned to the treatment group, irrespective of whether or not the household is enrolled in health insurance. This approach essentially aggregates all pre- and post-treatments periods, which is shown to reduce noise in the outcome variables and increase power (McKenzie, 2012).

To estimate the Average Treatment Effect on the Treated (ATT) rather than the ITT effects, we instrument whether the individual obtained health insurance in the post-intervention period with whether or not the individual lived in a treatment village, as follows.

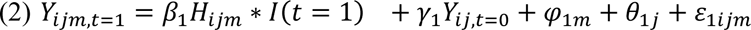

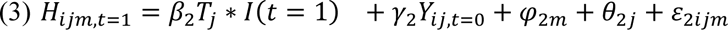

We also investigate heterogeneous effects by key baseline characteristics based on the program targeting criteria. To this end, the treatment group is interacted with the baseline subgroup indicators (or distance) as follows:

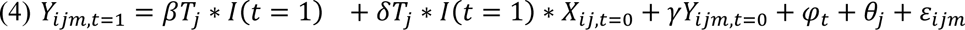

Heterogeneity is assessed by gender, age, baseline wealth (distinguishing between above and below median wealth, calculated as the first factor of a principal component analysis of dwelling characteristics and household assets), program-selected health facility (public versus private), and travel distance (measured as the distance between the household dwelling and the nearest i-PUSH-selected health facility).

## 4. RESULTS

### 4.1 Description of the study population

The demographic, socio-economic and health-related characteristics of the total analysis sample as measured in the baseline survey are shown in Table 1 column (1). Households had on average 4.9 members of whom 2.9 children below 18 years (and on average 1.3 children below age 5). The head of household was aged 37.1 years on average with 25.0 percent of heads being female. The vast majority of heads was married (91.4 percent) and 63.8 percent had completed primary education and above while the remaining one third had not completed primary schooling.

In 22 percent of households, at least one member was engaged in their own formal or informal business; in 9 percent of households someone worked on their own farm work; in 50 percent of households, at least one member engaged in casual labor; and in 21 percent of households, someone was engaged in wage work. Households reported a weekly income from work of on average KES 3,600 (averaged over months). At an average household size of almost five members, this suggests that the average individual in our study sample was living slightly below the poverty line^11^. Three quarters of households (76.3 percent) owned livestock and a similar proportion owned land (72.8 percent).

Households saved by keeping agricultural produce in stock, being member of a savings group or cooperation, or saving at other locations (at home, on M-PESA, or at a bank). Total household savings at baseline amounted to KES 12,276 on average. On the other hand, households had KES 3,885 in loans outstanding, either at a formal institution (22.3 percent) or from an informal lender (39.9 percent). They had also lent out money to others for an average of KES 1,892.

For the year prior to the baseline survey, households reported a total of 2.5 incidences of inpatient care, 15.3 cases of outpatient formal care, 14.5 visits to an informal drug vendor, 2.8 visits to a traditional health care provider, and 3.6 incidences of foregone care, i.e. indicating that a household member was ill but did not seek any care. Total reported household-level healthcare expenditures in the year prior to the survey were KES 8,134 on average.

### 4.2 Impact on health insurance enrolment

We start by investigating the impact of the program on enrolment in health insurance. The share of individuals who reported to be enrolled in NHIF increased steadily during program roll-out from June 2020 until October 2020 and reached a maximum of about 80 percent in the treatment group compared to a stable enrolment rate of a bit less than 20 percent in the control group (Figure 3, Panel a). At the household level, this translated into an enrolment rate of nearly 95 percent in the treatment group from October 2020 onwards (Figure 3, Panel b). These results suggest that the program was largely successful in enrolling the target population on NHIF.

**Figure 3.**
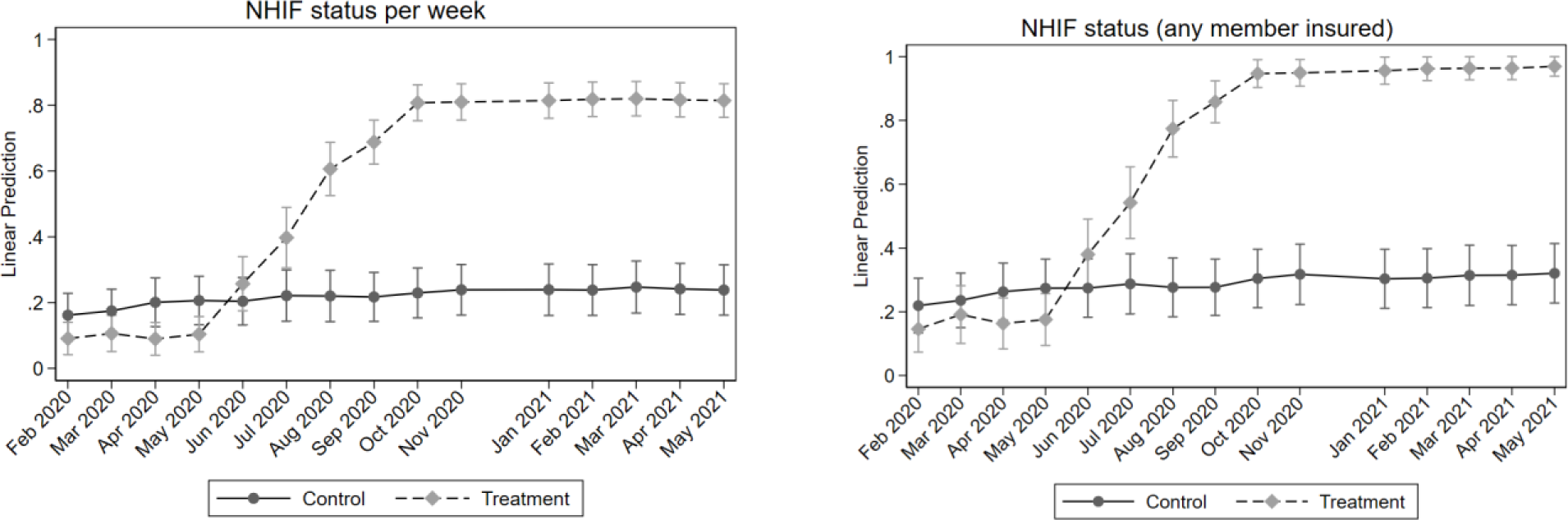
NHIF enrolment over time. Panel a): Share of individuals enrolled in NHIF per week, averaged over months; Panel b): Share of households with at least one member enrolled in NHIF per week, averaged over months

These descriptive results are confirmed in the impact regressions as shown in Table 2 Panel a). The ANCOVA ITT estimates (Column 2) indicate that the i-PUSH program increased the probability of being enrolled in NHIF with a significant 65.8 percentage points from a pre-intervention level in the control group of 18.9 percent (Column 1). When we look at the probability that *anyone* in the household is insured, the intervention brought the level from 25.3 percent pre-intervention in the control group to nearly universal with an increase of 71.4 percentage point.

**Table 2.**
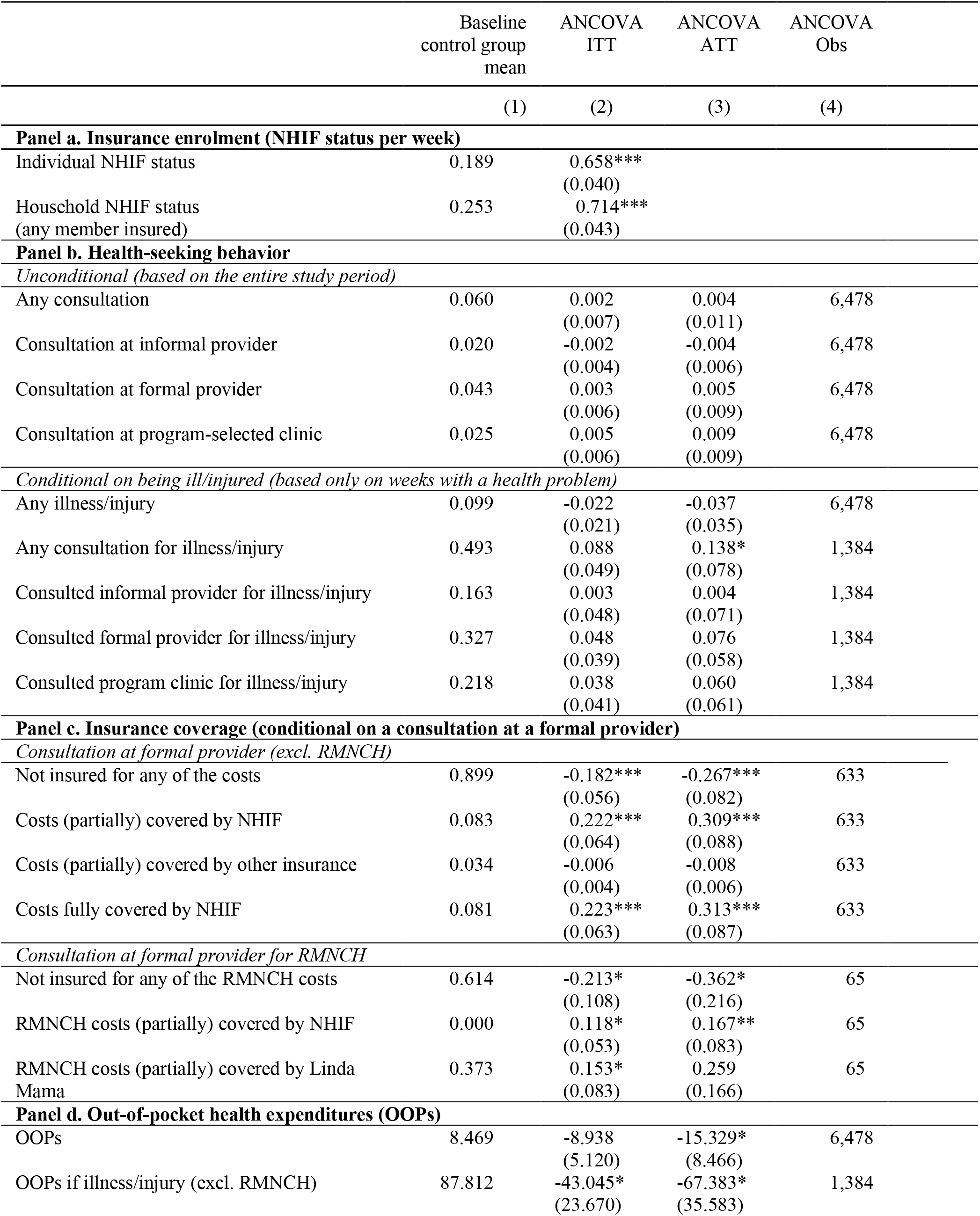
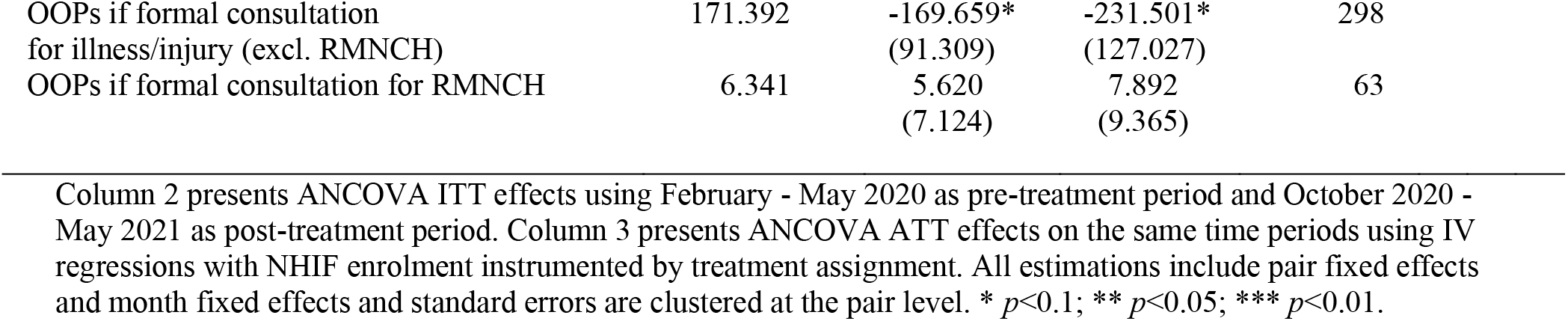
Program impact on key outcome variables.

The subgroup estimates based on the heterogeneity analyses are reported in Panel a) of the Appendix Tables A1-A7. We do not find a statistically significant difference in enrolment rates between adults and children (Table A1) nor between male and female adults (Table A2) or between boys and girls (Tables A3-A4).

Table A5 indicates that individuals in poor households (of below-median wealth) were less likely to be insured than individuals in non-poor households at baseline (11.2 vs 28.0 percent, respectively). The program reduced this gap with an estimated impact on the enrolment of poor and non-poor individuals of 70.5 and 59.9 percentage points, respectively. The differences between the impact estimates are not statistically significant, however.

We find a strong difference in the impact on enrolment between the study population linked to the private health facility compared to the public health facility (Table A6). Whereas their control baseline insurance rates were comparable at 20.7 and 18.3 percent, respectively, program impact is much larger for individuals living close to the private facility at 77.5 percentage points compared to the public facility at 62.9 percentage points, i.e. a 14.6 percentage points difference that is statistically significant.

One possible reason for the stronger impacts on insurance rates for households around the private facility may be related to its private ownership, with concomitantly higher costs. The public facility also reached NHIF level 4 at baseline, thereby increasing its fees as well; but that was not well-known yet around the time of enrolment, and hence may have attracted less enrollees. Another potential explanation may be related to differential perceptions of the quality of care, translating into lower willingness to enroll to benefit from public versus private care. Analyses of differential distances to the two providers suggest that travel considerations do not play a strong role in the decision to enroll in health insurance (Table A7).

### 4.3 Impact on healthcare utilization

The analysis of impact on health-seeking behavior starts with an investigation of *unconditional* healthcare utilization, i.e. the likelihood of any visit to a healthcare provider in any particular week. This analysis includes all individuals and all weeks from the entire study period in the estimations. In Table 2 Panel b) Column (1), we find that individuals in the control group visited a healthcare provider in 6.0 percent of all pre-intervention weeks (or conversely, in any pre-intervention week, on average 6.0 percent of control individuals visited a healthcare provider for any reason). They visited an informal provider in 2.0 percent, a formal provider (including program facilities) in 4.3 percent, and a program-selected provider in 2.5 percent of all pre-intervention weeks. The program did not significantly affect unconditional healthcare utilization at any type of provider for individuals living in the treatment communities (Column 2); that is, the Intention-to-Treat (ITT) effect is not significant. The Average Treatment Effect on the Treated (ATT), i.e. the effect on individuals in treatment communities who enrolled in insurance after the program was rolled out, is not significant either (Columns 3).

Heterogeneity analyses suggest that impacts are largest for individuals linked to the private rather than the public facility with estimates of 2.9 versus -0.2 percentage points, respectively, for the probability of visiting the program-selected clinic (Table A6). In line with these results, we find strong interaction effects with the distance to the clinic (Table A7). For individuals living very close next to a program-selected healthcare provider, consultations at a formal provider increase 2.5 percentage points from a baseline of 4.3 percent, fully driven by increased utilization of program clinics.

The next set of analyses in Panel b) investigates healthcare utilization, conditional on the individual experienced an illness or injury^12^. We have recorded 1,384 health problems in the health diaries. We start with an analysis of the observed trends in reporting an illness or injury. Pre-intervention, individuals in the control group reported a health problem in 9.9 percent of all weeks (Table 2 Column 1). Fluctuations in health problems over time are depicted in Figure 4. They are consistent with rain patterns and seasonal incidence of common infectious diseases such as malaria. Note that the peak in illnesses during the rainy season in 2020 was much lower than usual, most likely because of COVID-19 containment measures such as travel restrictions, social distancing, hand washing and so on (Gomez et al., 2023).

**Figure 4.**
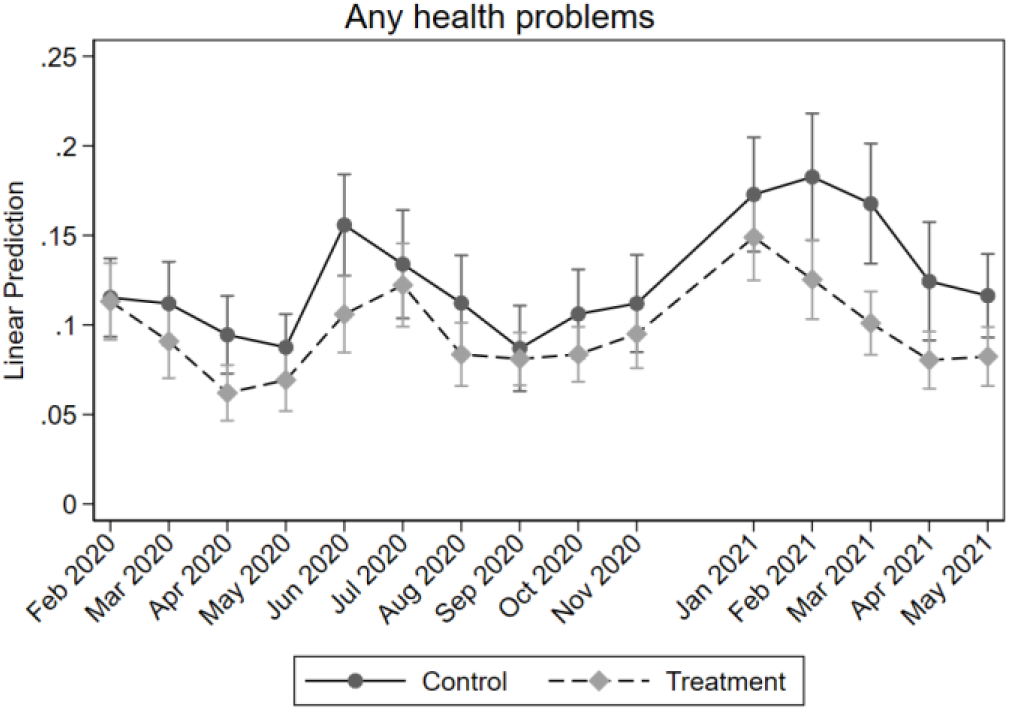
Occurrence of health problems (illness or injury) Note: Share of individuals reporting a health problem per week, averaged at the month level

The seasonal trends for the treatment and control groups overlap for most of the study period, except for two months pre-treatment (April and June 2020), and for four months towards the end of the period (February – May 2021), during which the treatment group reported significantly fewer health problems. A tentative explanation may be that these are also the peak months in terms of illnesses, increasing the power to statistically pick up improvements in health.

On average, however, the likelihood of a health shock did not change after the roll-out of the digital insurance scheme in the treatment villages, neither in the total sample (Table 2 Column 2) nor for individuals who actually enrolled in health insurance (Table 2 Column 3). Analyses by the various subgroups reveal little heterogeneity. For example, while male adults reported less health problems pre-intervention than female adults (7.8 versus 11.0 percent of weeks), the program did not differentially affect their health (Appendix Table A2). Boys and girls both of young and school-age report highly similar rates of illnesses and injuries pre- and post-intervention (Tables A3 and A4). Noteworthy is that individuals linked to the private program facility started off with less health issues at baseline compared to the public facility (in 8.9 vs. 10.1 percent of weeks), and they were more likely to be affected by the program roll-out with a significant increase in reported health shocks of 2.6 percentage points (Table A6), which might point to an increased awareness about illness and health among treated individuals.

We now turn to the impact estimates for healthcare utilization conditional on being ill or injured, reported in the lower part of Panel b). The impact estimates in Table 2 suggest that program impact on healthcare utilization was especially significant for individuals who actually enrolled in insurance. Their likelihood of seeking care at any type of provider increased with 13.8 percentage points from a pre-intervention control mean of 49.3 percent (ATT in Column 3). ITT impact estimates are also positive, but smaller in size and less precisely estimated. Pre-intervention, one-third of these consultations were at informal providers (for 16.3 percent of illness or injuries) and two-thirds (32.7 percent of health problems) were at formal providers. For 21.8 percent of health problems pre-intervention, individuals sought care at a program-linked healthcare provider. Although impact estimates for consultations at formal providers are statistically insignificant, they are positive and sizeable.

Figure 5 shows more detailed results over time: For most pre-intervention months, the treatment and control groups exhibit similar formal healthcare utilization rates conditional on being ill. From October 2020 onwards, when i-PUSH had been rolled out in all treatment villages, there is a significant increase in seeking care at formal providers for households in treatment communities compared to the control group.

**Figure 5.**
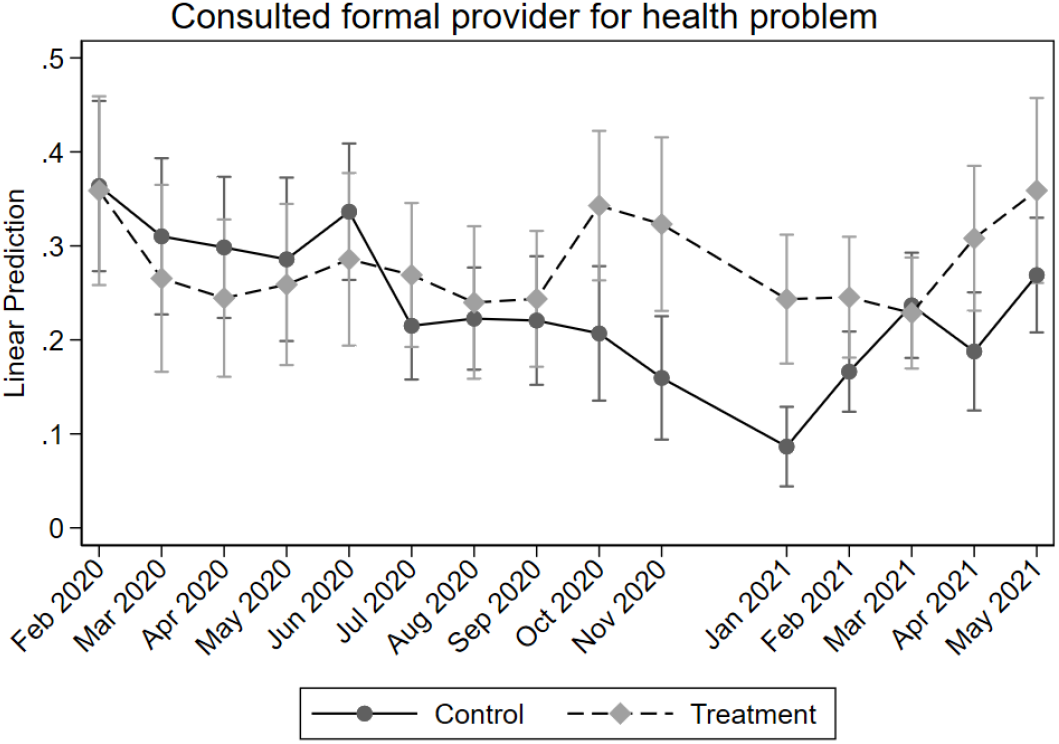
Formal health care consultation (conditional on illness/injury) Note: Share of individuals with a health problem who consulted a formal provider per week, averaged at the month level

Heterogeneity analyses show that the impact on healthcare utilization for a health problem is significantly larger for adult women at 13.6 percentage points compared to adult men at a non-significant -2.4, further increasing the pre-intervention gender gap in health-seeking behavior for health problems, which was 48.4 percent versus 39.2 percent for women versus men, respectively (Table A2). The impact on adult women is mostly driven by an increase in visits to program-selected providers, although individual impact estimates are not significant at conventional levels.

Gender differences among children are less pronounced (Tables A3-A4). School-aged boys (5-18 years) in treatment communities are more likely to consult a healthcare provider as a result of the program, but this concerns mostly *informal* care, while informal care for girls decreases – especially for girls below 5.

The impact of the program on seeking care at a program-selected provider again varies significantly by linked facility (Table A6). At baseline in the control group, individuals in the catchment area of the private facility sought care at this provider only for 8.3 percent of their illnesses or injuries, compared to 25.7 percent of individuals in the catchment area of the public facility. The program increased conditional healthcare utilization at the private provider with 17.4 percentage points while it did not significantly increase visits to the public provider.

Note that the overall effect on seeking care when ill on utilization is greater in communities linked to the public hospital. In this area the program resulted in an increase in conditional health-seeking behavior of 12.9 percent. However, none of this effect was realized in the public facility itself (the point estimate is slightly negative). The estimates suggest that most of the increased utilization was at formal health care providers not selected by the insurance program. This is in line with the fact that the choice set of potential NHIF healthcare providers was greater in that area, including the two unselected facilities.

The impact is substantially larger for those who live close to the program-selected facility (Table A7). For those who live next to their linked facility, the program increases the probability of seeking care at a formal provider when ill with a significant 15.0 percentage points. For every kilometer greater distance, the effect reduces by 6.4 percentage points. The pattern is almost identical when we focus on seeking care at a program-linked facility. The impact of the digital insurance scheme is thus very sensitive to how accessible the selected healthcare providers are.

Distance may be one of the drivers why the impacts are generally stronger for households linked to the private facility as compared to households linked to the public facility: part of the households linked to the public facility were located in the catchment area of the two private healthcare providers that were ultimately not included in the program; thus on average, these households live further from the program-selected clinic.

### 4.4 Impact on financial coverage of medical costs

This section investigates to what extent the increased enrolment in health translates into greater coverage of health-related costs at formal providers and lower out-of-pocket expenditures for health care. We start with the impact on financial coverage (Table 2 Panel C). This panel focuses on consultations at formal providers, as only those are potentially covered by health insurance. We distinguish between consultations for RMNCH care (including family planning, ANC, delivery, PNC, childhood immunization and health check-ups/monitoring of children below 5) in the bottom rows and any other consultations in the top rows. In our analysis dataset, there are n=878 RMNCH consultations and n= 6,115 other (non-RMNCH) consultations, of which n=4,800 are directly related to an acute illness or injury and n=1,104 are for drug supplies (mostly for longer-term or chronic health conditions). Note that these numbers represent individual consultations, whereas Table 2 aggregates the data to the individual-month level.

At baseline in the control communities, individuals were not insured for any of their medical costs in 89.9 percent of their formal consultations. This percentage decreased significantly for individuals in the treatment group after program roll-out with 18.2 percentage points, and 26.7 percentage points for those that actually enrolled in insurance. Conversely, while only 8.3 percent of control individuals pre-intervention had some or all of their costs covered by NHIF, the program had a significant positive impact on financial coverage of 22.2 percentage points on average and 30.9 percentage points for insured individuals. Pre-intervention, 3.4 percent of formal consultations were covered by other schemes; this did not change after program roll-out.

Similarly, we find that 61.4 percent of pre-intervention RMNCH consultations were not covered by any scheme, and the digital insurance program reduced this with about one third, or 21.3 percentage points on average (ITT) and 36.2 percentage points for those insured (ATT). Pre-intervention, RMNCH consultations were covered 37.3 percent of the time by Linda Mama and never by NHIF. Interestingly, the program increased Linda Mama coverage with 15.3 percentage points for individuals in treatment villages, perhaps through greater awareness and knowledge of this scheme. The impact on financial coverage was 11.8 percentage points at the overall village level and 16.7 percentage points for insured individuals.

As shown in the Appendix Tables, impact on financial coverage of general healthcare utilization is significantly larger for adult women than adult men: Whereas treated women see an increase in partially or fully covered costs of 26.8 percentage points, the impact for men is not discernable from zero (Table A2). The differential impact on school-aged girls versus boys is even larger at 45.2 and 11.2 percentages points, respectively (Table A3). This might be related to the greater need for healthcare among adolescent girls who enter their reproductive years. We do not find any differential impact on financial coverage for girls versus boys below 5 (Table A4).

Impacts on financial coverage are also largest for individuals linked to the private rather than the public health facility, with an increase in medical costs partially or fully covered by NHIF of 37.2 and 16.6 percentage points, respectively (Table A6). This can be explained by the joint effect of both a larger impact on insurance enrolment and a greater increase in visits to the private program provider. As for other outcomes, we find no differential impact for adults versus children (Table A1) or wealth (Table A5).

### 4.5 Impact on out-of-pocket expenditures (OOPs)

Financially, we investigate program impact on out-of-pocket health expenditures. The level of weekly health expenditures (averaged on a monthly basis for easy visualization) is presented in Figure 6. The left-hand panel shows unconditional OOPs (i.e. regardless of the occurrence of a health problem in a particular week), while the right-hand panel shows OOPs only in weeks when the individual was ill or injured. We observe a diverging trend from September 2020 onwards, after which the OOPs in the control group are always above the treatment group expenditures. These differences are statistically significant for the months of February and March 2021.

**Figure 6.**
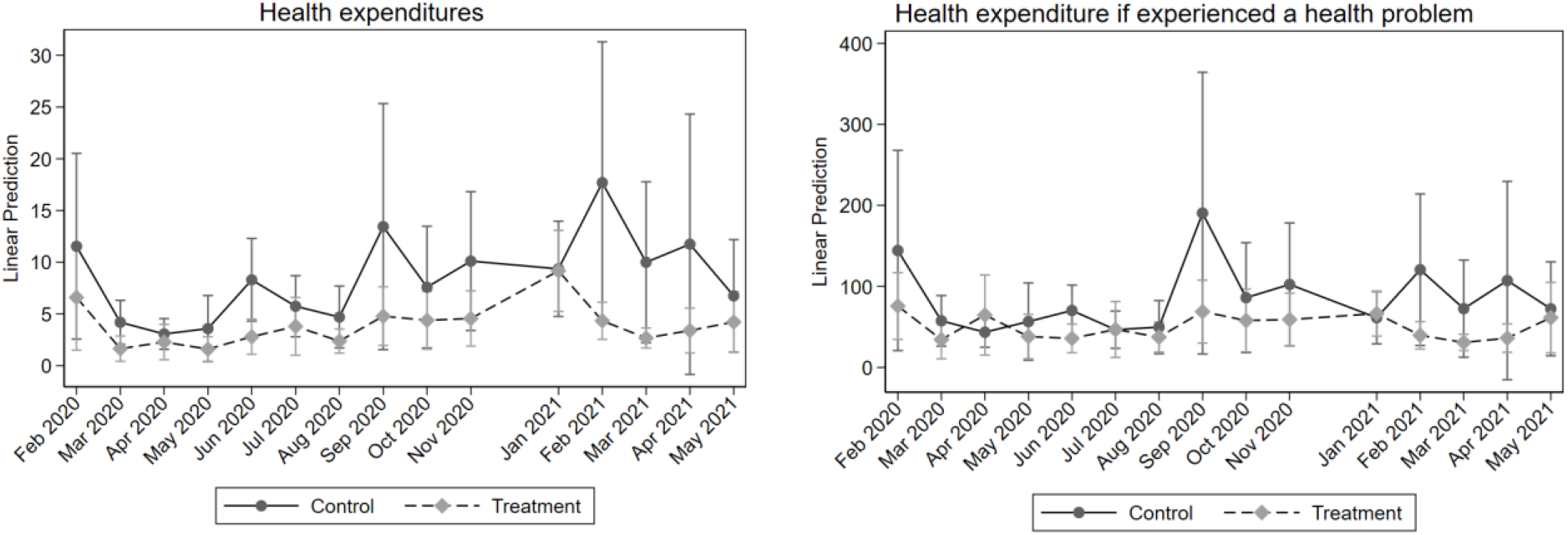
Individual OOPs unconditional and conditional on being ill.

Table 2 Panel d) confirms that the program significantly reduced individual-level OOPs. Column 1 shows that, pre-intervention, individuals in control communities spent on average KES 8.47 per week (independent of their health status and health care utilization), and KES 440 per year. Individuals in treatment villages saw their average weekly health expenditures drop by KES 8.94 per week on average as a result of the program, translating into a reduction in annual OOPs of KES 465. Focusing only on the treatment individuals who actually took insurance, we find that the program facilitated an overall weekly (annual) reduction in unconditional OOPs of KES 15.33 (KES 797), so independent of whether an individual was ill or not during the year. We emphasize that these are average values, which may mask substantial variation in the study sample, especially when many individuals have low expenditures while a few have high OOPs.

We now zoom in on OOPs only for those weeks in which individuals were experiencing a health problem. The pre-intervention average of OOPs in such weeks was KES 87.81 for control individuals. The program reduced these OOPs by more than half (by KES 43.05 for the average individual in treatment villages, and by KES 67.38 for insured individuals in treatment villages). Impacts are substantially larger when focusing only on those occurrences of health problems for which individuals sought formal care. Columns (2) and (3) show that the digital insurance scheme is estimated to reduce OOPs in weeks with a formal consultation (excluding RMNCH consultations) with KES 169.66 on average in treatment villages, and with KES 231.50 for insured individuals in treatment villages. No effects were found for OOPs related to RMNCH care, which were very low to start with. Appendix Table A8 breaks down the OOPs impact estimates by sub-category of health spending, including consultation fees, registration, drugs, laboratory and transport costs, and other costs. The results indicate that the largest savings due to the insurance scheme are on spending for medication.

We do not find differential impacts on OOPs by gender or age (Tables A1-A4). Impact estimates between the lower and the higher wealth groups are not statistically significant either (Table A5), although the point estimates suggest that wealthier individuals benefited more from the program. This may be explained by their much larger baseline OOPs compared to the poorest half of individuals. Finally, we find that OOPs conditional on having a health problem reduced most for individuals linked to the private rather than the public facility (Table A6). This differential drop in OOPs is not linked to travel distance, but rather seems driven by a greater likelihood of individuals to use their new NHIF card at the private facility.

Table 3 translates the individual OOPs into monthly household-level OOPs and compares them to households’ monthly capacity to pay (CTP) to estimate the likelihood of catastrophic health expenditures (CHE). As shown, the average weekly OOPs aggregated to the household-month level significantly decreased due to the program. Households’ CTP on the other hand, did not change significantly for treated versus control households. The household OOPs as a share of CTP were 11.0 percent at baseline in the control group, which decreased with 5.9 percentage points for households in the treatment communities and 8.7 percentage points for treated households who took insurance. As a result, the probability of incurring CHE (at the 40% of CTP threshold) dropped sharply with 7.4 percentage points (ITT) from a likelihood of 10.1 percent at baseline.

**Table 3.**
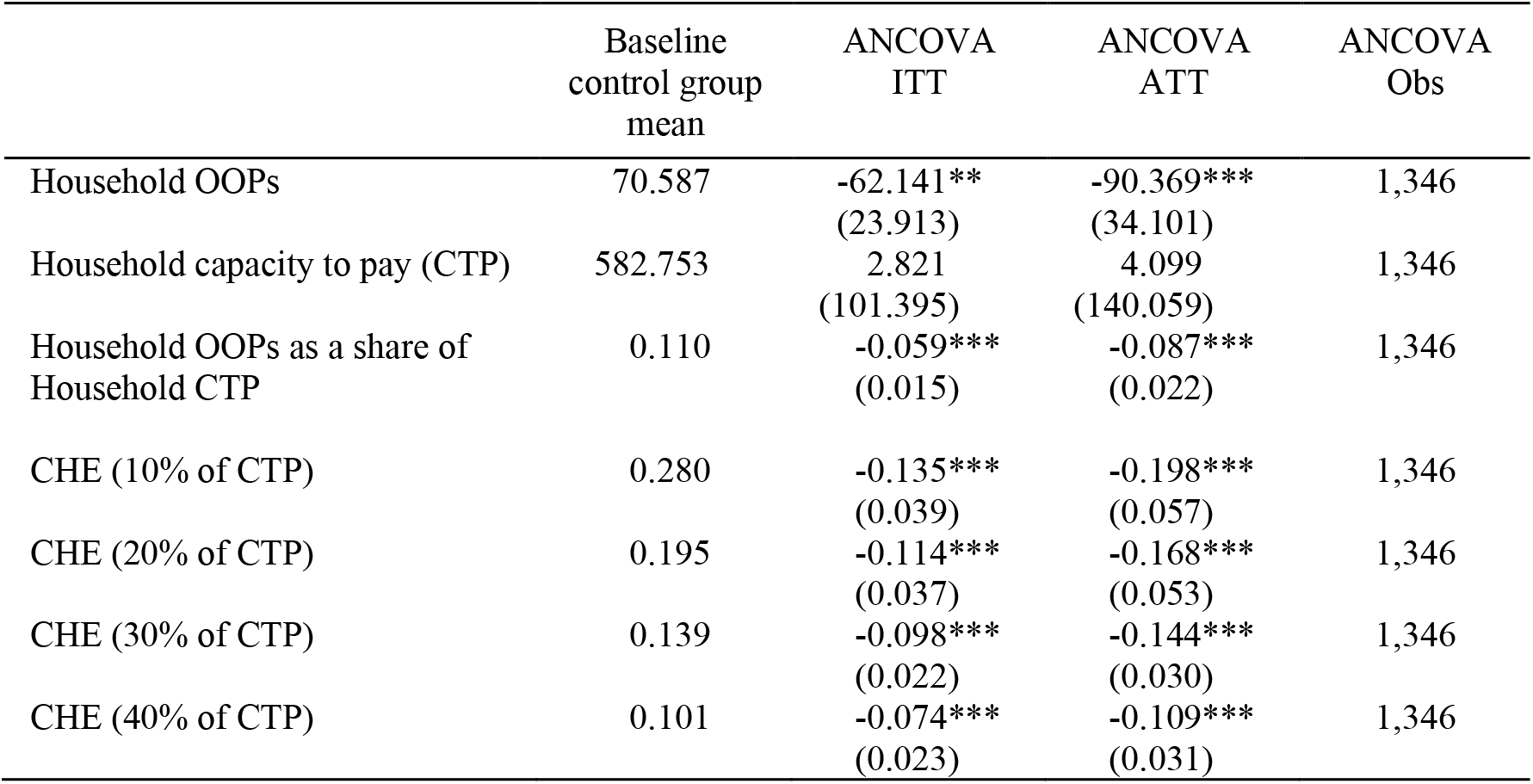
Impact on catastrophic health expenditures (CHE) at the household-month level.

|  | Baseline<br>control group<br>mean | ANCOVA<br>ITT | ANCOVA<br>ATT | ANCOVA<br>Obs |
| --- | --- | --- | --- | --- |
| Household OOPs | 70.587 | -62.141**<br>(23.913) | -90.369***<br>(34.101) | 1,346 |
| Household capacity to pay (CTP) | 582.753 | 2.821<br>(101.395) | 4.099<br>(140.059) | 1,346 |
| Household OOPs as a share of<br>Household CTP | 0.110 | -0.059***<br>(0.015) | -0.087***<br>(0.022) | 1,346 |
| CHE (10% of CTP) | 0.280 | -0.135***<br>(0.039) | -0.198***<br>(0.057) | 1,346 |
| CHE (20% of CTP) | 0.195 | -0.114***<br>(0.037) | -0.168***<br>(0.053) | 1,346 |
| CHE (30% of CTP) | 0.139 | -0.098***<br>(0.022) | -0.144***<br>(0.030) | 1,346 |
| CHE (40% of CTP) | 0.101 | -0.074***<br>(0.023) | -0.109***<br>(0.031) | 1,346 |

## 5. DISCUSSION AND CONCLUSION

This study evaluated a subsidized digital health insurance program in Western Kenya. The program aimed to enhance access to quality healthcare for low-income women of reproductive age and their family members. It alleviated women’s financial constraints to formal healthcare utilization by offering subsidized, mobile phone-based health insurance, while upgrading selected healthcare providers, enhancing the training of community health workers and improving decision-making capacity of local policymakers through better data on healthcare utilization. By targeting women directly rather than their husbands, the program aimed to empower women in their ability to access healthcare for themselves, their children, and their spouses.

Our findings show that the program was highly successful in enrolling target households (i.e. households including at least one woman of reproductive age who was either pregnant or with a child below age four living with her) on the NHIF insurance scheme. Impact on individual insurance uptake was 65.8 percentage points (from a baseline control mean 18.9 percent), while the impact on household insurance uptake (i.e. households that had enrolled at least one household member) was 71.4 percentage points compared to a baseline control mean of 25.3 percent, reaching almost full household coverage in the treatment group. This success stands in sharp contrast to a large recent experiment in Indonesia that also provided a full subsidy for health insurance and assistance in registration, which increased enrolment by only 24 percent point from a base of 8 percent; 54 percent made an attempt to enroll in the Indonesian scheme, but less than half succeeded to do so (Banerjee et al., 2021).

The Kenyan program not only reduced costs of enrolment, but also worked towards eliminating various other (administrative, logistical, and trust) barriers to enrolment. The introduction of the program and its agents by trusted local CHWs, the hands-on assistance with the digital registration procedures at women’s homes, and support in retrieving the necessary documentation such as children’s birth certificates, likely have all contributed to the high enrolment rates.

We note, however, that 9% of the target women in the original sample did not have a national ID-card – a general NHIF requirement for registration. Their households were excluded from the study and replaced. These are likely to be among the most disadvantaged households in the study area, suggesting that administrative requirements of national insurance programs that aim at the poor may miss out precisely on those families that would benefit most. In addition, many children did not have the required birth certificates to be included on their mother’s cover. With substantial efforts, program staff supported these families in obtaining the certificates, raising awareness along the way among government and NHIF officials about such registration hurdles. Currently, the Kakamega Registrar has taken up these challenges and the County is moving towards an easier and faster procedure to request birth certificates for children.

Despite the full premium subsidy, a non-negligible proportion of about 20 percent of household members in the treatment group did not enroll. In general, we find that enrolment rates are slightly higher for women than for men, and slightly higher for adults than for children, albeit not significantly so. The inability to provide the required birth certificates for some children, and a national ID-card for husbands who are often away from home for longer periods of time, may have contributed to these (small) differences in enrolment.

The less than full enrolment at the household level (i.e. the cases in which the entire household declined to enroll) may be explained by the list of NHIF-empaneled providers: Households were significantly more likely to enroll when they could select a private facility as their preferred NHIF provider compared to households who were matched to a public NHIF facility. This is not necessarily due to a lower perceived quality of care at the public hospital, since a sizeable share of respondents initially mentioned this facility as their preferred provider. However, a substantial number of households were living in the catchment areas of other facilities that were ultimately dropped from the program. They were encouraged to choose the public facility instead, which was further away, and which may have contributed to their reluctance to enroll.

Unfortunately, our data do not allow us to investigate NHIF renewal rates. By the endline in June 2021, the program in the study area had ended and both the premium subsidy as well as the ease of having digital access to healthcare were stopped. Several initiatives are currently ongoing in collaboration with Kakamega County officials to investigate how the improvements in UHC can be further sustained.

Impacts on healthcare utilization were more ambiguous. Consultations at program-selected and upgraded providers did not consistently increase, neither unconditional (curative plus preventive and related to RMNCH) or conditional on having a health problem (curative care only). However, this masks substantial geographical heterogeneity. The impact on visiting a program provider for treatment households linked to the private facility was large and significant at 2.9 percentage points per week on average, while for the households linked to the public hospital there was no significant effect on average. This can be explained to a large extent by the larger travel distances of study households linked to the public facility; indeed, treatment households living next to their linked facility were 2.5 percentage points more likely to visit a formal provider compared to control households (from a baseline control mean of 4.3 percent), while every additional kilometer between an individual’s dwelling and the i-PUSH provider reduced program impact on the likelihood of provider consultations with 1.4 percentage points. This highlights the importance of geographical accessibility and the need to have enough NHIF-empaneled providers also in more remote, rural areas in order to enhance UHC.

Conditional on illness, those who took up health insurance utilized health services more often. Their likelihood of seeking care at any type of provider increased with 13.8 percentage points from a pre-intervention control mean of 49.3 percent. The increase in utilization materialized from October 2020 onwards, when the roll-out of the program in the treatment communities (i.e. the sign up of interested households on the digital insurance scheme) had been completed. The effects are stronger for adult women than for adult men, in line with program expectations.

The high rates of insurance enrolment for treatment households translated into a significant improvement in financial coverage of medical costs. The program produced a significant increase in the financial coverage of illnesses and injuries by NHIF of 22.2 percentage points (compared to 8.3 percent in the control group at baseline) and 30.9 percentage points when focusing solely on treatment households who actually enrolled in NHIF. Overall, the program reduced the average health expenditures for individual household members living in treatment villages with 8.94 KES per week (or 465 KES on an annual basis), and with 15.33 KES weekly (or 797 KES annually) when focusing on insured individuals in the treatment area. Unexpectedly, the program also increased coverage of RMNCH costs, both through the NHIF cover but also through Linda Mama – the latter perhaps driven by increased awareness of the scheme in treatment villages. These reductions in health spending in turn substantially enhanced households’ financial protection: the likelihood of incurring catastrophic health expenditures reduced substantially in treatment villages and was virtually eliminated for treatment households who enrolled in the digital scheme.

To conclude, the mobile phone-based, subsidized health insurance coverage was successful in increasing health insurance enrollment and reducing out-of-pocket expenditures for low-income households in rural Western Kenya. When living sufficiently close to one of the program-selected clinics (that had participated in the quality improvement program SafeCare), women in treatment villages were significantly more likely to seek health care for their children and for themselves at a formal provider.

Follow-up analysis with the financial and health diaries data will allow for an improved understanding of informal risk-coping mechanisms with and without formal insurance, such as informal gift-giving and loans, (dis)saving, asset sales or adjustments in hours worked. Further qualitative and behavioral research may shed light on the specific role that digital technology may have played in yielding these impacts. In particular, to what extent did the fact that women could get the insurance card on their own sim-card (rather than their husbands’) stimulate their uptake of insurance, and utilization of the scheme? Finally, a willingness-to-pay (WTP) experiment conducted at the endline survey will yield further insights into households’ willingness to enroll in health insurance at different premium levels, and show whether a one-year experience with insurance coverage influences WTP.

## Data Availability

All data produced in the present study will be deposited in an online repository in the near future and will be available upon reasonable request to the authors.

## Acknowledgements

We greatly appreciate the continued support and many fruitful discussions with PharmAccess Foundation and Amref Health Africa and their i-PUSH program staff, and the continued high-quality data collection efforts of the field research team in Khwisero. Our study participants, who offered us their valuable time and provided so many insights into their lives, deserve our special gratitude. This work is funded by the Dutch National Postcode Lottery, the Joep Lange Institute, and the Dutch Ministry of Foreign Affairs through the Health Insurance Fund.

## Appendix A

**Figure A1.**
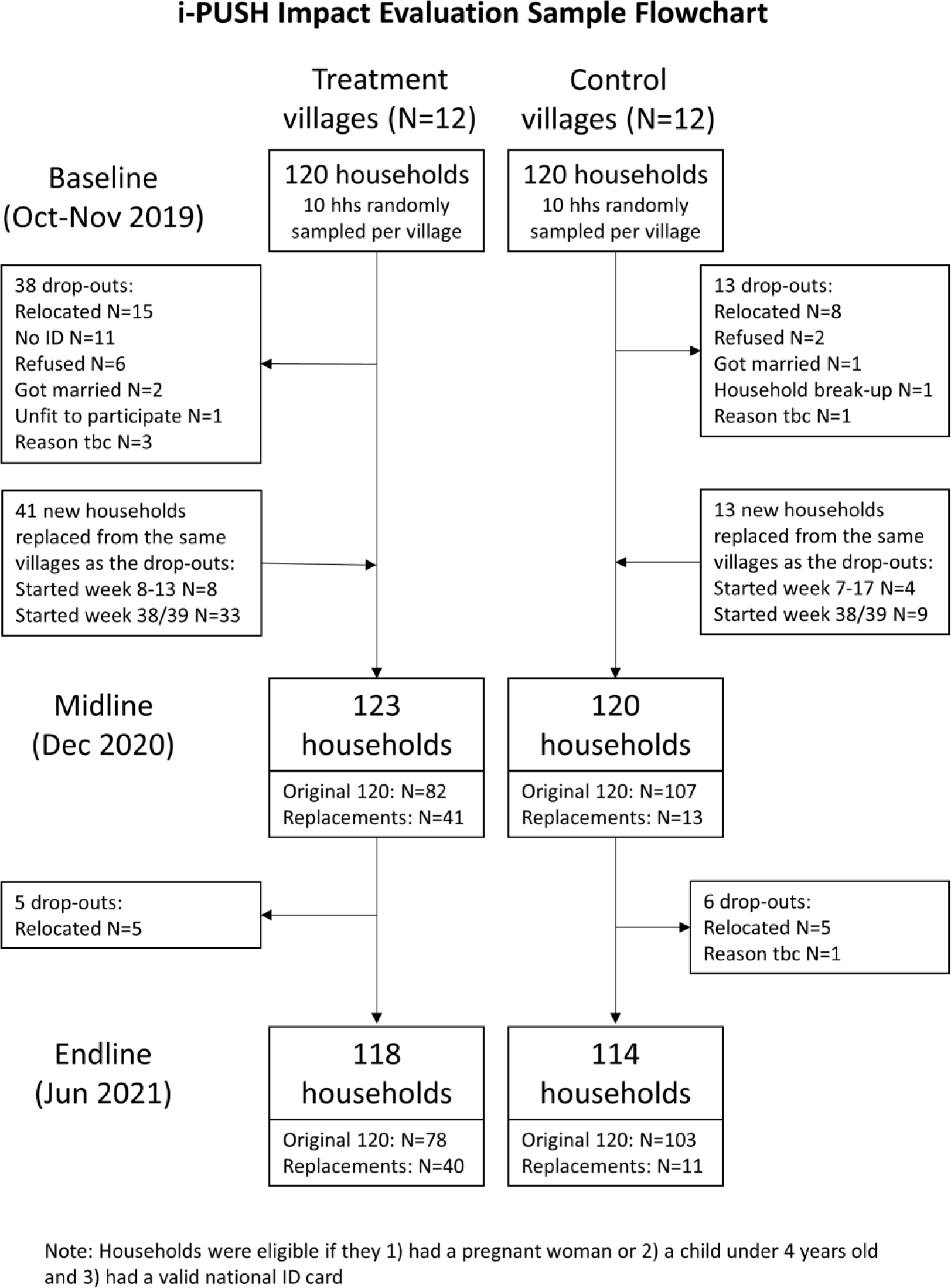
Sample flowchart.

**Table A1.** Heterogeneous impact estimates – by age: adults (18 years+) vs. children (< 18)

|  | Baseline control group<br>mean |  | Impact estimate |  | P-value of<br>difference | Obs |
| --- | --- | --- | --- | --- | --- | --- |
|  | 18 years<br>or older | Under 18<br>years | 18 years<br>or older | Under 18<br>years |  |  |
| **Panel a. Insurance enrolment (NHIF status per week)** |  |  |  |  |  |  |
| Individual NHIF status | 0.191 | 0.187 | 0.700***<br>(0.044) | 0.633***<br>(0.046) | 0.156 | 6,478 |
| Household NHIF status<br>(any member insured) | 0.229 | 0.271 | 0.708***<br>(0.044) | 0.718***<br>(0.045) | 0.650 | 6,478 |
| **Panel b. Health-seeking behavior** |  |  |  |  |  |  |
| *Unconditional (based on the entire study period)* |  |  |  |  |  |  |
| Any health problem (per<br>week) | 0.097 | 0.100 | -0.024<br>(0.025) | -0.021<br>(0.021) | 0.880 | 6,478 |
| Any consultation (per<br>week) | 0.054 | 0.064 | 0.003<br>(0.011) | 0.002<br>(0.008) | 0.923 | 6,478 |
| Any consultation at<br>Informal provider (per wk) | 0.020 | 0.021 | -0.001<br>(0.007) | -0.003<br>(0.007) | 0.876 | 6,478 |
| Any consultation at<br>formal provider (per week) | 0.037 | 0.047 | 0.003<br>(0.008) | 0.003<br>(0.005) | 0.947 | 6,478 |
| Any consultation at<br>i-PUSH clinic (per week) | 0.022 | 0.027 | 0.006<br>(0.005) | 0.005<br>(0.006) | 0.821 | 6,478 |
| *Conditional on being ill/injured (based only on weeks with a health problem)* |  |  |  |  |  |  |
| Any consultation for<br>health problem | 0.451 | 0.522 | 0.065<br>(0.063) | 0.102<br>(0.068) | 0.677 | 1,384 |
| Consulted informal provider<br>for health problem | 0.187 | 0.148 | 0.011<br>(0.049) | -0.003<br>(0.073) | 0.881 | 1,384 |
| Consulted formal provider<br>for health problem | 0.262 | 0.370 | 0.038<br>(0.059) | 0.054<br>(0.038) | 0.783 | 1,384 |
| Consulted i-PUSH clinic<br>for health problem | 0.189 | 0.237 | 0.053<br>(0.041) | 0.026<br>(0.045) | 0.430 | 1,384 |
| **Panel c. Insurance coverage (conditional on a consultation at a formal provider)** |  |  |  |  |  |  |
| *Consultation at formal provider for health problem (excl. MNCH)* |  |  |  |  |  |  |
| Not insured for any of the<br>costs | 0.873 | 0.915 | -0.165**<br>(0.062) | -0.194**<br>(0.078) | 0.764 | 633 |
| Costs (partially) covered by<br>NHIF/M-TIBA | 0.101 | 0.072 | 0.212***<br>(0.059) | 0.229**<br>(0.088) | 0.856 | 633 |
| Costs (partially) covered by<br>other insurance | 0.053 | 0.023 | -0.014<br>(0.010) | 0.000<br>(0.001) | 0.198 | 633 |
| Costs fully covered by<br>NHIF/M-TIBA | 0.096 | 0.072 | 0.201***<br>(0.062) | 0.238**<br>(0.088) | 0.718 | 633 |
| **Panel d. Out-of-pocket health expenditures (OOPs)** |  |  |  |  |  |  |
| OOPs | 10.008 | 7.347 | -5.661<br>(6.861) | -10.652**<br>(4.351) | 0.252 | 6,478 |
| OOPs if health problem | 105.940 | 75.765 | -16.862<br>(28.714) | -58.588*<br>(26.993) | 0.210 | 1,384 |
| OOPs if formal consultation<br>for health problem (excl.<br>MNCH) | 248.099 | 135.294 | -63.149<br>(82.693) | -236.471*<br>(124.617) | 0.221 | 298 |

The estimates present ANCOVA ITT effects using February - May 2020 as pre-treatment period and October 2020 - May 2021 as post-treatment period. In these estimations, the treatment dummy is interacted with a dummy for being under 18 to obtain pint estimates for both subgroups and the p-value of the difference between sub-groups. All estimations include pair fixed effects and month fixed effects and standard errors are clustered at the pair level. \* $p < 0.1$ ; \*\* $p < 0.05$ ; \*\*\* $p < 0.01$ .

**Table A2.** Heterogeneous impact estimates - by gender (for adults)

|  | Baseline control group<br>mean |  | Impact estimate |  | P-value of<br>difference | Obs |
| --- | --- | --- | --- | --- | --- | --- |
|  | Male<br>adults | Female<br>adults | Male<br>adults | Female<br>adults |  |  |
| **Panel a. Insurance enrolment (NHIF status per week)** |  |  |  |  |  |  |
| Individual NHIF status | 0.181 | 0.198 | 0.624***<br>(0.065) | 0.738***<br>(0.045) | 0.099* | 2,427 |
| Household NHIF status<br>(any member insured) | 0.225 | 0.231 | 0.720***<br>(0.050) | 0.698***<br>(0.047) | 0.598 | 2,427 |
| **Panel b. Health-seeking behavior** |  |  |  |  |  |  |
| *Unconditional (based on the entire study period)* |  |  |  |  |  |  |
| Any health problem (per<br>week) | 0.078 | 0.110 | -0.025<br>(0.026) | -0.025<br>(0.025) | 0.997 | 2,427 |
| Any consultation (per<br>week) | 0.031 | 0.071 | -0.004<br>(0.012) | 0.006<br>(0.010) | 0.162 | 2,427 |
| Any consultation at<br>Informal provider (per wk) | 0.017 | 0.022 | -0.001<br>(0.008) | -0.001<br>(0.006) | 0.970 | 2,427 |
| Any consultation at<br>formal provider (per week) | 0.015 | 0.053 | -0.003<br>(0.007) | 0.007<br>(0.009) | 0.108 | 2,427 |
| Any consultation at<br>i-PUSH clinic (per week) | 0.008 | 0.032 | 0.007**<br>(0.003) | 0.004<br>(0.007) | 0.515 | 2,427 |
| *Conditional on being ill/injured (based only on weeks with a health problem)* |  |  |  |  |  |  |
| Any consultation for<br>health problem | 0.392 | 0.484 | -0.024<br>(0.100) | 0.136**<br>(0.061) | 0.089* | 543 |
| Consulted informal provider<br>for health problem | 0.224 | 0.166 | 0.065<br>(0.077) | -0.012<br>(0.051) | 0.390 | 543 |
| Consulted formal provider<br>for health problem | 0.168 | 0.314 | -0.066<br>(0.083) | 0.107<br>(0.061) | 0.020** | 543 |
| Consulted i-PUSH clinic<br>for health problem | 0.156 | 0.208 | -0.014<br>(0.047) | 0.094<br>(0.054) | 0.166 | 543 |
| **Panel c. Insurance coverage (conditional on a consultation at a formal provider)** |  |  |  |  |  |  |
| *Consultation at formal provider for health problem (excl. MNCH)* |  |  |  |  |  |  |
| Not insured for any of the<br>costs | 0.926 | 0.848 | 0.022<br>(0.084) | -0.254**<br>(0.082) | 0.042** | 249 |
| Costs (partially) covered by<br>NHIF/M-TIBA | 0.083 | 0.109 | 0.009<br>(0.093) | 0.268***<br>(0.070) | 0.037** | 249 |
| Costs (partially) covered by<br>other insurance | 0.000 | 0.077 | 0.002<br>(0.005) | -0.023<br>(0.019) | 0.194 | 249 |
| Costs fully covered by<br>NHIF/M-TIBA | 0.083 | 0.103 | -0.027<br>(0.102) | 0.261***<br>(0.073) | 0.038** | 249 |
| **Panel d. Out-of-pocket health expenditures (OOPs)** |  |  |  |  |  |  |
| OOPs | 9.020 | 10.722 | -5.564<br>(7.240) | -5.127<br>(7.310) | 0.942 | 2,427 |
| OOPs if health problem | 96.644 | 111.105 | -42.997<br>(47.284) | 2.882<br>(40.096) | 0.491 | 543 |
| OOPs if formal consultation<br>for health problem (excl.<br>MNCH) | 326.333 | 224.150 | -422.707<br>(426.852) | -86.215<br>(62.079) | 0.460 | 108 |

The estimates present ANCOVA ITT effects using February - May 2020 as pre-treatment period and October 2020 - May 2021 as post-treatment period. In these estimations, the treatment dummy is interacted with a dummy for being female to obtain point estimates for both subgroups and the p-value of the difference between subgroups. The sample is restricted to the adult population 18 years and older. All estimations include pair fixed effects and month fixed effects and standard errors are clustered at the pair level. \* $p < 0.1$ ; \*\* $p < 0.05$ ; \*\*\* $p < 0.01$ .

**Table A3.** Heterogeneous impact estimates - by gender (for children 5-18 years)

|  | Baseline control group<br>mean |  | Impact estimate |  | P-value of<br>difference | Obs |
| --- | --- | --- | --- | --- | --- | --- |
|  | Boys 5-18<br>years | Girls 5-18<br>years | Boys 5-18<br>years | Girls 5-18<br>years |  |  |
| **Panel a. Insurance enrolment (NHIF status per week)** |  |  |  |  |  |  |
| Individual NHIF status | 0.227 | 0.238 | 0.648***<br>(0.067) | 0.650***<br>(0.063) | 0.982 | 2,299 |
| Household NHIF status<br>(any member insured) | 0.285 | 0.303 | 0.692***<br>(0.070) | 0.755***<br>(0.056) | 0.349 | 2,299 |
| **Panel b. Health-seeking behavior** |  |  |  |  |  |  |
| *Unconditional (based on the entire study period)* |  |  |  |  |  |  |
| Any health problem (per<br>week) | 0.067 | 0.068 | -0.017<br>(0.016) | -0.013<br>(0.019) | 0.852 | 2,299 |
| Any consultation (per<br>week) | 0.031 | 0.046 | 0.008<br>(0.009) | 0.001<br>(0.009) | 0.333 | 2,299 |
| Any consultation at<br>Informal provider (per wk) | 0.010 | 0.016 | 0.007<br>(0.007) | -0.008<br>(0.007) | 0.092* | 2,299 |
| Any consultation at<br>formal provider (per week) | 0.022 | 0.034 | 0.001<br>(0.007) | 0.007<br>(0.005) | 0.282 | 2,299 |
| Any consultation at<br>i-PUSH clinic (per week) | 0.017 | 0.018 | 0.001<br>(0.007) | 0.003<br>(0.005) | 0.596 | 2,299 |
| *Conditional on being ill/injured (based only on weeks with a health problem)* |  |  |  |  |  |  |
| Any consultation for<br>health problem | 0.443 | 0.611 | 0.333**<br>(0.142) | 0.093<br>(0.102) | 0.184 | 235 |
| Consulted informal provider<br>for health problem | 0.144 | 0.149 | 0.235<br>(0.167) | 0.027<br>(0.097) | 0.280 | 235 |
| Consulted formal provider<br>for health problem | 0.322 | 0.470 | 0.040<br>(0.131) | 0.062<br>(0.121) | 0.903 | 235 |
| Consulted i-PUSH clinic<br>for health problem | 0.287 | 0.232 | -0.065<br>(0.065) | 0.005<br>(0.124) | 0.677 | 235 |
| **Panel c. Insurance coverage (conditional on a consultation at a formal provider)** |  |  |  |  |  |  |
| *Consultation at formal provider for health problem (excl. MNCH)* |  |  |  |  |  |  |
| Not insured for any of the<br>costs | 1.000 | 0.864 | -0.145<br>(0.144) | -0.500**<br>(0.166) | 0.098* | 105 |
| Costs (partially) covered by<br>NHIF/M-TIBA | 0.000 | 0.091 | 0.112<br>(0.142) | 0.452**<br>(0.171) | 0.095* | 105 |
| Costs (partially) covered by<br>other insurance | 0.000 | 0.045 | 0.000<br>(0.000) | 0.000<br>(0.000) | . | 105 |
| Costs fully covered by<br>NHIF/M-TIBA | 0.000 | 0.091 | 0.101<br>(0.147) | 0.460**<br>(0.163) | 0.085* | 105 |
| **Panel d. Out-of-pocket health expenditures (OOPs)** |  |  |  |  |  |  |
| OOPs | 3.848 | 3.579 | -2.419<br>(2.835) | -9.502<br>(5.619) | 0.305 | 2,299 |
| OOPs if health problem | 59.713 | 37.350 | 50.622*<br>(27.623) | -42.904<br>(38.509) | 0.133 | 235 |
| OOPs if formal consultation<br>for health problem (excl.<br>MNCH) | 86.364 | 37.656 | 1,205.338<br>(865.441) | -1,032.663<br>(656.068) | 0.161 | 53 |

The estimates present ANCOVA ITT effects using February - May 2020 as pre-treatment period and October 2020 - May 2021 as post-treatment period. In these estimations, the treatment dummy is interacted with a dummy for being female to obtain point estimates for both subgroups and the p-value of the difference between subgroups. The sample is restricted to the child population aged 5-18 years. All estimations include pair fixed effects and month fixed effects and standard errors are clustered at the pair level. \* $p < 0.1$ ; \*\* $p < 0.05$ ; \*\*\* $p < 0.01$ .

**Table A4.** Heterogeneous impact estimates - by gender (for children under 5 years)

|  | Baseline control group<br>mean |  | Impact estimate |  | P-value of<br>difference | Obs |
| --- | --- | --- | --- | --- | --- | --- |
|  | Boys<br>under 5<br>years | Girls<br>under 5<br>years | Boys<br>under 5<br>years | Girls<br>under 5<br>years |  |  |
| **Panel a. Insurance enrolment (NHIF status per week)** |  |  |  |  |  |  |
| Individual NHIF status | 0.132 | 0.136 | 0.654***<br>(0.073) | 0.594***<br>(0.064) | 0.546 | 1,813 |
| Household NHIF status<br>(any member insured) | 0.230 | 0.257 | 0.696***<br>(0.065) | 0.721***<br>(0.045) | 0.732 | 1,813 |
| **Panel b. Health-seeking behavior** |  |  |  |  |  |  |
| *Unconditional (based on the entire study period)* |  |  |  |  |  |  |
| Any health problem (per<br>week) | 0.135 | 0.138 | -0.035<br>(0.039) | -0.021<br>(0.034) | 0.692 | 1,813 |
| Any consultation (per<br>week) | 0.097 | 0.095 | 0.013<br>(0.018) | -0.016<br>(0.013) | 0.149 | 1,813 |
| Any consultation at<br>Informal provider (per wk) | 0.030 | 0.026 | -0.003<br>(0.014) | -0.008<br>(0.008) | 0.729 | 1,813 |
| Any consultation at<br>formal provider (per week) | 0.074 | 0.072 | 0.014<br>(0.012) | -0.012<br>(0.011) | 0.082* | 1,813 |
| Any consultation at<br>i-PUSH clinic (per week) | 0.047 | 0.035 | 0.023*<br>(0.013) | -0.003<br>(0.008) | 0.043 | 1,813 |
| *Conditional on being ill/injured (based only on weeks with a health problem)* |  |  |  |  |  |  |
| Any consultation for<br>health problem | 0.548 | 0.499 | 0.158<br>(0.112) | -0.051<br>(0.065) | 0.068* | 611 |
| Consulted informal provider<br>for health problem | 0.159 | 0.128 | 0.036<br>(0.108) | -0.127**<br>(0.048) | 0.075* | 611 |
| Consulted formal provider<br>for health problem | 0.389 | 0.334 | 0.059<br>(0.048) | 0.032<br>(0.081) | 0.758 | 611 |
| Consulted i-PUSH clinic<br>for health problem | 0.260 | 0.199 | 0.087<br>(0.063) | -0.004<br>(0.056) | 0.180 | 611 |
| **Panel c. Insurance coverage (conditional on a consultation at a formal provider)** |  |  |  |  |  |  |
| *Consultation at formal provider for health problem (excl. MNCH)* |  |  |  |  |  |  |
| Not insured for any of the<br>costs | 0.877 | 0.930 | -0.152<br>(0.125) | -0.241**<br>(0.101) | 0.332 | 283 |
| Costs (partially) covered by<br>NHIF/M-TIBA | 0.123 | 0.020 | 0.201<br>(0.130) | 0.242**<br>(0.110) | 0.640 | 283 |
| Costs (partially) covered by<br>other insurance | 0.027 | 0.050 | 0.000<br>(0.000) | 0.000<br>(0.000) | . | 283 |
| Costs fully covered by<br>NHIF/M-TIBA | 0.123 | 0.020 | 0.202<br>(0.130) | 0.251**<br>(0.107) | 0.561 | 283 |
| **Panel d. Out-of-pocket health expenditures (OOPs)** |  |  |  |  |  |  |
| OOPs | 5.416 | 19.837 | -23.879**<br>(9.485) | -6.050**<br>(2.678) | 0.052* | 1,813 |
| OOPs if health problem | 39.474 | 157.471 | -85.948<br>(54.194) | -50.091*<br>(24.463) | 0.513 | 611 |
| OOPs if formal consultation<br>for health problem (excl. MNCH) | 25.490 | 399.857 | -504.324<br>(373.157) | 14.903<br>(47.301) | 0.186 | 141 |

The estimates present ANCOVA ITT effects using February - May 2020 as pre-treatment period and October 2020 - May 2021 as post-treatment period. In these estimations, the treatment dummy is interacted with a dummy for being female to obtain point estimates for both subgroups and the p-value of the difference between subgroups. The sample is restricted to the child population aged under 5 years. All estimations include pair fixed effects and month fixed effects and standard errors are clustered at the pair level. \* $p < 0.1$ ; \*\* $p < 0.05$ ; \*\*\* $p < 0.01$ .

**Table A5.** Heterogeneous impact estimates - by wealth of the household.

|  | Baseline control group<br>mean |  | Impact estimate |  | P-value of<br>difference | Obs |
| --- | --- | --- | --- | --- | --- | --- |
|  | Low<br>wealth | High<br>wealth | Low<br>wealth | High<br>wealth |  |  |
| **Panel a. Insurance enrolment (NHIF status per week)** |  |  |  |  |  |  |
| Individual NHIF status | 0.112 | 0.280 | 0.705***<br>(0.063) | 0.599***<br>(0.050) | 0.216 | 6,478 |
| Household NHIF status<br>(any member insured) | 0.140 | 0.388 | 0.773***<br>(0.067) | 0.642***<br>(0.061) | 0.192 | 6,478 |
| **Panel b. Health-seeking behavior** |  |  |  |  |  |  |
| *Unconditional (based on the entire study period)* |  |  |  |  |  |  |
| Any health problem (per<br>week) | 0.099 | 0.098 | -0.034<br>(0.026) | -0.008<br>(0.020) | 0.254 | 6,478 |
| Any consultation (per<br>week) | 0.059 | 0.061 | -0.001<br>(0.007) | 0.005<br>(0.011) | 0.680 | 6,478 |
| Any consultation at<br>Informal provider (per wk) | 0.017 | 0.024 | -0.004<br>(0.005) | -0.000<br>(0.005) | 0.573 | 6,478 |
| Any consultation at<br>formal provider (per week) | 0.045 | 0.041 | 0.001<br>(0.007) | 0.004<br>(0.010) | 0.795 | 6,478 |
| Any consultation at<br>i-PUSH clinic (per week) | 0.028 | 0.022 | 0.003<br>(0.006) | 0.008<br>(0.009) | 0.599 | 6,478 |
| *Conditional on being ill/injured (based only on weeks with a health problem)* |  |  |  |  |  |  |
| Any consultation for<br>health problem | 0.474 | 0.516 | 0.111<br>(0.072) | 0.048<br>(0.058) | 0.499 | 1,384 |
| Consulted informal provider<br>for health problem | 0.133 | 0.200 | 0.021<br>(0.066) | -0.022<br>(0.039) | 0.475 | 1,384 |
| Consulted formal provider<br>for health problem | 0.345 | 0.304 | 0.052<br>(0.035) | 0.032<br>(0.069) | 0.802 | 1,384 |
| Consulted i-PUSH clinic<br>for health problem | 0.243 | 0.187 | 0.075<br>(0.045) | -0.004<br>(0.073) | 0.392 | 1,384 |
| **Panel c. Insurance coverage (conditional on a consultation at a formal provider)** |  |  |  |  |  |  |
| *Consultation at formal provider for health problem (excl. MNCH)* |  |  |  |  |  |  |
| Not insured for any of the<br>costs | 0.901 | 0.898 | -0.132<br>(0.076) | -0.208***<br>(0.058) | 0.449 | 633 |
| Costs (partially) covered by<br>NHIF/M-TIBA | 0.077 | 0.089 | 0.185*<br>(0.095) | 0.233***<br>(0.063) | 0.658 | 633 |
| Costs (partially) covered by<br>other insurance | 0.030 | 0.039 | -0.011<br>(0.007) | 0.000<br>(0.003) | 0.218 | 633 |
| Costs fully covered by<br>NHIF/M-TIBA | 0.074 | 0.089 | 0.185*<br>(0.094) | 0.236***<br>(0.059) | 0.626 | 633 |
| **Panel d. Out-of-pocket health expenditures (OOPs)** |  |  |  |  |  |  |
| OOPs | 4.071 | 13.712 | -0.906<br>(3.094) | -18.737<br>(11.356) | 0.193 | 6,478 |
| OOPs if health problem | 34.734 | 152.041 | -9.988<br>(33.742) | -89.081*<br>(40.738) | 0.205 | 1,384 |
| OOPs if formal consultation<br>for health problem (excl.<br>MNCH) | 42.304 | 346.039 | -183.827<br>(154.432) | -139.590**<br>(61.063) | 0.742 | 298 |

The estimates present ANCOVA ITT effects using February - May 2020 as pre-treatment period and October 2020 - May 2021 as post-treatment period. In these estimations, the treatment dummy is interacted with a dummy for being above the median of the wealth index to obtain point estimates for both subgroups and the p-value of the difference between subgroups. All estimations include pair fixed effects and month fixed effects and standard errors are clustered at the pair level. \* $p < 0.1$ ; \*\* $p < 0.05$ ; \*\*\* $p < 0.01$ .

**Table A6.** Heterogeneous impact estimates - by program clinic.

|  | Baseline control group<br>mean |  | Impact estimate |  | P-value of<br>difference | Obs |
| --- | --- | --- | --- | --- | --- | --- |
|  | private | public | private | public |  |  |
| **Panel a. Insurance enrolment (NHIF status per week)** |  |  |  |  |  |  |
| Individual NHIF status | 0.207 | 0.183 | 0.761***<br>(0.041) | 0.624***<br>(0.044) | 0.037** | 6,478 |
| Household NHIF status<br>(any member insured) | 0.270 | 0.248 | 0.799***<br>(0.059) | 0.687***<br>(0.049) | 0.157 | 6,478 |
| **Panel b. Health-seeking behavior** |  |  |  |  |  |  |
| *Unconditional (based on the entire study period)* |  |  |  |  |  |  |
| Any health problem (per<br>week) | 0.089 | 0.101 | 0.026*<br>(0.013) | -0.038<br>(0.025) | 0.050* | 6,478 |
| Any consultation (per<br>week) | 0.069 | 0.057 | 0.007<br>(0.016) | 0.001<br>(0.007) | 0.714 | 6,478 |
| Any consultation at<br>Informal provider (per wk) | 0.025 | 0.019 | -0.006<br>(0.005) | -0.001<br>(0.005) | 0.457 | 6,478 |
| Any consultation at<br>formal provider (per week) | 0.047 | 0.042 | 0.009<br>(0.012) | 0.001<br>(0.006) | 0.561 | 6,478 |
| Any consultation at<br>i-PUSH clinic (per week) | 0.020 | 0.026 | 0.029**<br>(0.013) | -0.002<br>(0.003) | 0.038** | 6,478 |
| *Conditional on being ill/injured (based only on weeks with a health problem)* |  |  |  |  |  |  |
| Any consultation for<br>health problem | 0.520 | 0.486 | -0.032<br>(0.048) | 0.129**<br>(0.055) | 0.050* | 1,384 |
| Consulted informal provider<br>for health problem | 0.229 | 0.144 | -0.075<br>(0.055) | 0.029<br>(0.056) | 0.215 | 1,384 |
| Consulted formal provider<br>for health problem | 0.291 | 0.337 | 0.005<br>(0.043) | 0.064<br>(0.048) | 0.379 | 1,384 |
| Consulted i-PUSH clinic<br>for health problem | 0.083 | 0.257 | 0.174**<br>(0.068) | -0.010<br>(0.033) | 0.031** | 1,384 |
| **Panel c. Insurance coverage (conditional on a consultation at a formal provider)** |  |  |  |  |  |  |
| *Consultation at formal provider for health problem (excl. MNCH)* |  |  |  |  |  |  |
| Not insured for any of the<br>costs | 0.875 | 0.907 | -0.276***<br>(0.055) | -0.146*<br>(0.072) | 0.184 | 633 |
| Costs (partially) covered by<br>NHIF/M-TIBA | 0.097 | 0.078 | 0.372***<br>(0.039) | 0.166**<br>(0.075) | 0.038** | 633 |
| Costs (partially) covered by<br>other insurance | 0.042 | 0.032 | 0.002<br>(0.002) | -0.008<br>(0.006) | 0.220 | 633 |
| Costs fully covered by<br>NHIF/M-TIBA | 0.097 | 0.076 | 0.365***<br>(0.031) | 0.170**<br>(0.074) | 0.037** | 633 |
| **Panel d. Out-of-pocket health expenditures (OOPs)** |  |  |  |  |  |  |
| OOPs | 6.539 | 9.068 | -10.369**<br>(4.114) | -8.464<br>(6.711) | 0.815 | 6,478 |
| OOPs if health problem | 66.427 | 93.997 | -111.958**<br>(43.835) | -19.247<br>(24.541) | 0.096* | 1,384 |
| OOPs if formal consultation<br>for health problem (excl.<br>MNCH) | 53.902 | 201.688 | -293.423<br>(188.207) | -110.886<br>(96.896) | 0.404 | 298 |

The estimates present ANCOVA ITT effects using February - May 2020 as pre-treatment period and October 2020 - May 2021 as post-treatment period. In these estimations, the treatment dummy is interacted with a dummy for being linked to Khisero HC to obtain point estimates for both subgroups and the p-value of the difference between subgroups. All estimations include pair fixed effects and month fixed effects and standard errors are clustered at the pair level. \* $p < 0.1$ ; \*\* $p < 0.05$ ; \*\*\* $p < 0.01$ .

**Table A7.** Heterogeneous impact estimates - by distance to program clinic.

|  | Baseline control<br>group mean | Main effect | Interaction with<br>distance | Obs |
| --- | --- | --- | --- | --- |
| **Panel a. Insurance enrolment (NHIF status per week)** |  |  |  |  |
| Individual NHIF status | 0.189 | 0.715***<br>(0.076) | -0.033<br>(0.039) | 6,478 |
| Household NHIF status<br>(any member insured) | 0.253 | 0.748***<br>(0.084) | -0.020<br>(0.042) | 6,478 |
| **Panel b. Health-seeking behavior** |  |  |  |  |
| *Unconditional (based on the entire study period)* |  |  |  |  |
| Any health problem (per<br>week) | 0.099 | 0.039<br>(0.027) | -0.036**<br>(0.015) | 6,478 |
| Any consultation (per<br>week) | 0.060 | 0.029**<br>(0.013) | -0.016**<br>(0.005) | 6,478 |
| Any consultation at<br>Informal provider (per wk) | 0.020 | 0.004<br>(0.008) | -0.004<br>(0.004) | 6,478 |
| Any consultation at<br>formal provider (per week) | 0.043 | 0.025**<br>(0.009) | -0.014***<br>(0.004) | 6,478 |
| Any consultation at<br>i-PUSH clinic (per week) | 0.025 | 0.027**<br>(0.010) | -0.013***<br>(0.004) | 6,478 |
| *Conditional on being ill/injured (based only on weeks with a health problem)* |  |  |  |  |
| Any consultation for<br>health problem | 0.493 | 0.077<br>(0.076) | 0.003<br>(0.047) | 1,384 |
| Consulted informal provider<br>for health problem | 0.163 | -0.069<br>(0.059) | 0.042<br>(0.037) | 1,384 |
| Consulted formal provider<br>for health problem | 0.327 | 0.150***<br>(0.046) | -0.064**<br>(0.026) | 1,384 |
| Consulted i-PUSH clinic<br>for health problem | 0.218 | 0.144<br>(0.083) | -0.066*<br>(0.035) | 1,384 |
| **Panel c. Insurance coverage (conditional on a consultation at a formal provider)** |  |  |  |  |
| *Consultation at formal provider for health problem (excl. MNCH)* |  |  |  |  |
| Not insured for any of the<br>costs | 0.899 | -0.302***<br>(0.053) | 0.076**<br>(0.030) | 633 |
| Costs (partially) covered by<br>NHIF/M-TIBA | 0.083 | 0.388***<br>(0.053) | -0.105***<br>(0.032) | 633 |
| Costs (partially) covered by<br>other insurance | 0.034 | -0.008<br>(0.007) | 0.001<br>(0.002) | 633 |
| Costs fully covered by<br>NHIF/M-TIBA | 0.081 | 0.391***<br>(0.049) | -0.107***<br>(0.033) | 633 |
| **Panel d. Out-of-pocket health expenditures (OOPs)** |  |  |  |  |
| OOPs | 8.469 | -7.719<br>(5.796) | -0.492<br>(1.820) | 6,478 |
| OOPs if health problem | 87.812 | -114.768<br>(64.869) | 47.374<br>(31.212) | 1,384 |
| OOPs if formal consultation for<br>health problem (excl. MNCH) | 171.392 | -328.777<br>(201.225) | 105.086<br>(77.754) | 298 |

The estimates present ANCOVA ITT effects using February - May 2020 as pre-treatment period and October 2020 - May 2021 as post-treatment period. In these estimations, the treatment dummy is interacted with the distance in km to either Khwisero (if linked to Khwisero) or Namasoli (if linked to Namasoli). Also a dummy for being linked to Namasoli or Khwisero is included. All estimations include pair fixed effects and month fixed effects and standard errors are clustered at the pair level. \* $p < 0.1$ ; \*\* $p < 0.05$ ; \*\*\* $p < 0.01$ .

**Table A8.**
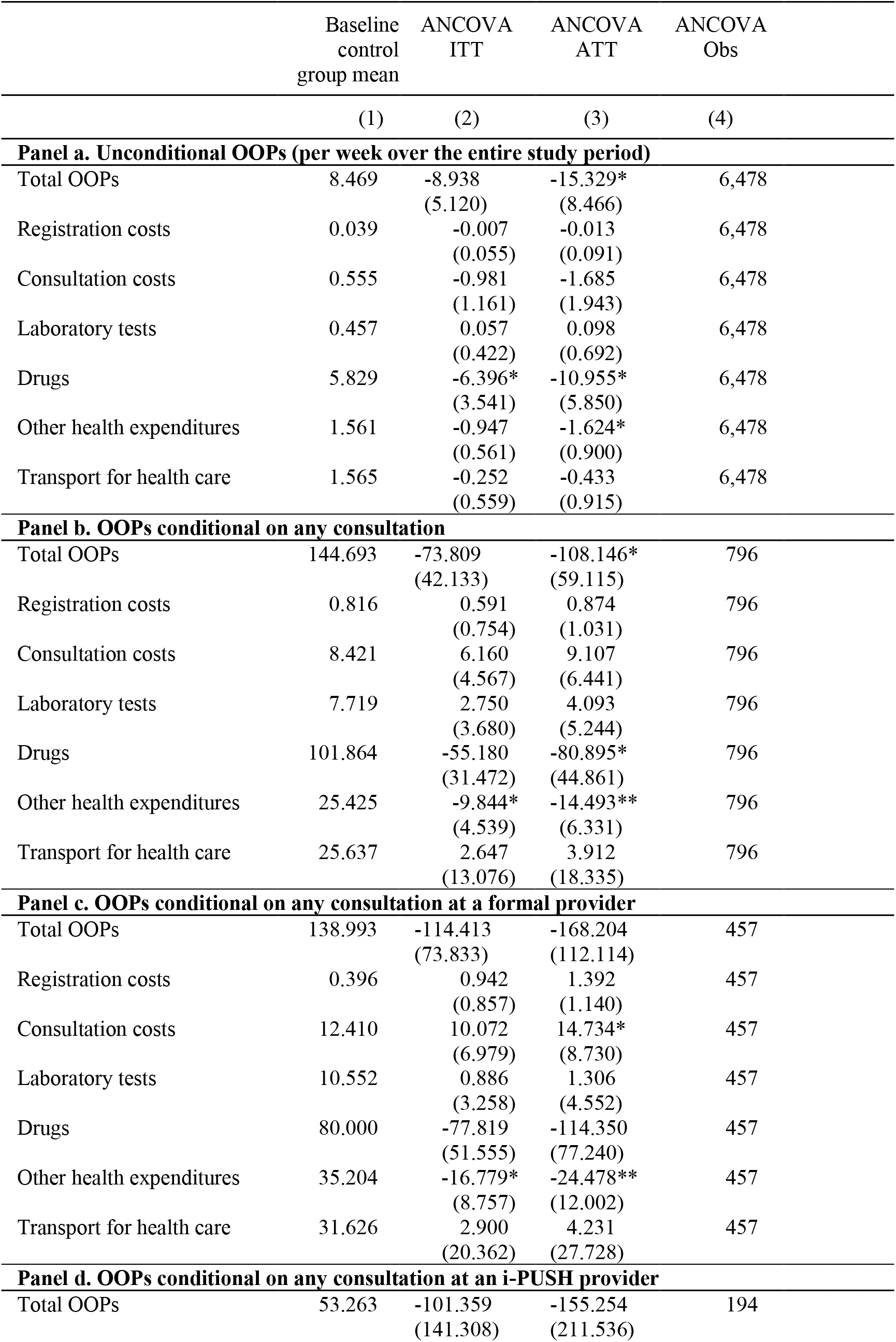
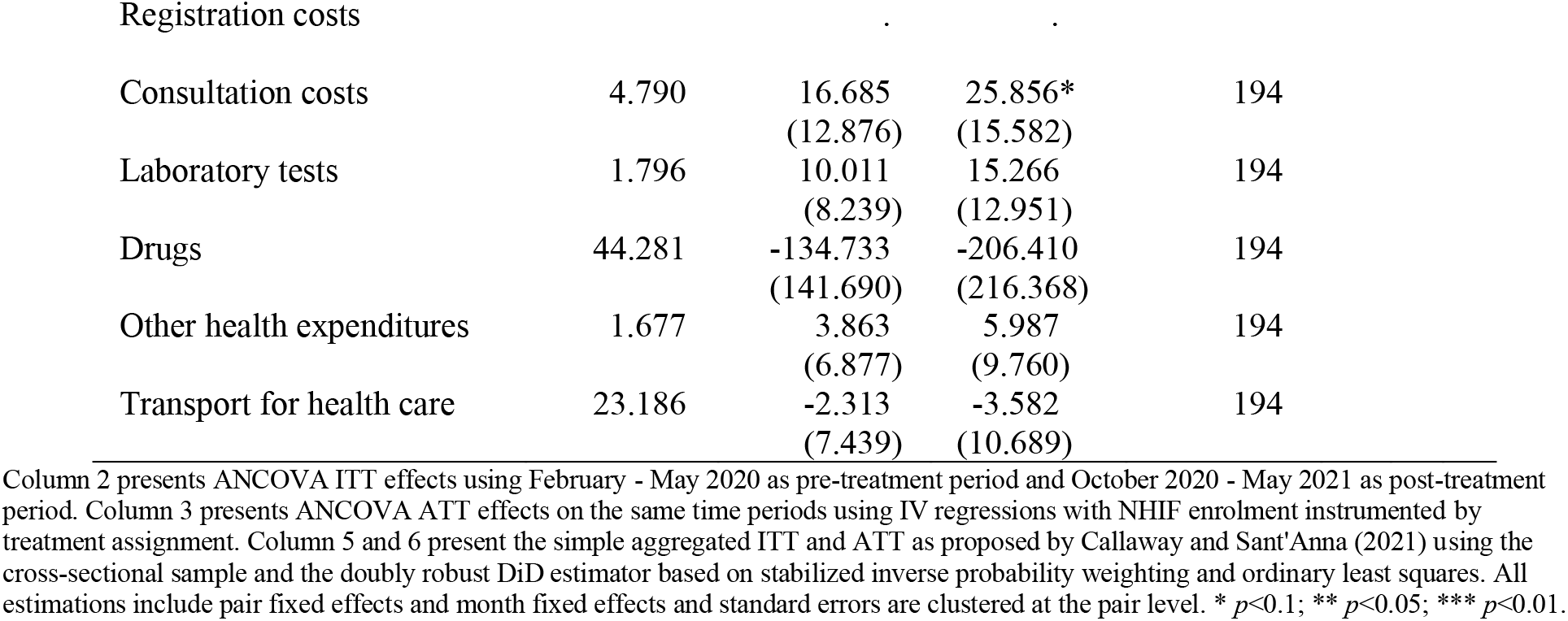
Impact on out-of-pocket health expenditures (OOPs) by type of costs.

|  | Baseline<br>control<br>group mean | ANCOVA<br>ITT | ANCOVA<br>ATT | ANCOVA<br>Obs |
| --- | --- | --- | --- | --- |
|  | (1) | (2) | (3) | (4) |
| **Panel a. Unconditional OOPs (per week over the entire study period)** |  |  |  |  |
| Total OOPs | 8.469 | -8.938<br>(5.120) | -15.329*<br>(8.466) | 6,478 |
| Registration costs | 0.039 | -0.007<br>(0.055) | -0.013<br>(0.091) | 6,478 |
| Consultation costs | 0.555 | -0.981<br>(1.161) | -1.685<br>(1.943) | 6,478 |
| Laboratory tests | 0.457 | 0.057<br>(0.422) | 0.098<br>(0.692) | 6,478 |
| Drugs | 5.829 | -6.396*<br>(3.541) | -10.955*<br>(5.850) | 6,478 |
| Other health expenditures | 1.561 | -0.947<br>(0.561) | -1.624*<br>(0.900) | 6,478 |
| Transport for health care | 1.565 | -0.252<br>(0.559) | -0.433<br>(0.915) | 6,478 |
| **Panel b. OOPs conditional on any consultation** |  |  |  |  |
| Total OOPs | 144.693 | -73.809<br>(42.133) | -108.146*<br>(59.115) | 796 |
| Registration costs | 0.816 | 0.591<br>(0.754) | 0.874<br>(1.031) | 796 |
| Consultation costs | 8.421 | 6.160<br>(4.567) | 9.107<br>(6.441) | 796 |
| Laboratory tests | 7.719 | 2.750<br>(3.680) | 4.093<br>(5.244) | 796 |
| Drugs | 101.864 | -55.180<br>(31.472) | -80.895*<br>(44.861) | 796 |
| Other health expenditures | 25.425 | -9.844*<br>(4.539) | -14.493**<br>(6.331) | 796 |
| Transport for health care | 25.637 | 2.647<br>(13.076) | 3.912<br>(18.335) | 796 |
| **Panel c. OOPs conditional on any consultation at a formal provider** |  |  |  |  |
| Total OOPs | 138.993 | -114.413<br>(73.833) | -168.204<br>(112.114) | 457 |
| Registration costs | 0.396 | 0.942<br>(0.857) | 1.392<br>(1.140) | 457 |
| Consultation costs | 12.410 | 10.072<br>(6.979) | 14.734*<br>(8.730) | 457 |
| Laboratory tests | 10.552 | 0.886<br>(3.258) | 1.306<br>(4.552) | 457 |
| Drugs | 80.000 | -77.819<br>(51.555) | -114.350<br>(77.240) | 457 |
| Other health expenditures | 35.204 | -16.779*<br>(8.757) | -24.478**<br>(12.002) | 457 |
| Transport for health care | 31.626 | 2.900<br>(20.362) | 4.231<br>(27.728) | 457 |
| **Panel d. OOPs conditional on any consultation at an i-PUSH provider** |  |  |  |  |
| Total OOPs | 53.263 | -101.359<br>(141.308) | -155.254<br>(211.536) | 194 |
| Registration costs |  | . | . |  |
| Consultation costs | 4.790 | 16.685<br>(12.876) | 25.856*<br>(15.582) | 194 |
| Laboratory tests | 1.796 | 10.011<br>(8.239) | 15.266<br>(12.951) | 194 |
| Drugs | 44.281 | -134.733<br>(141.690) | -206.410<br>(216.368) | 194 |
| Other health expenditures | 1.677 | 3.863<br>(6.877) | 5.987<br>(9.760) | 194 |
| Transport for health care | 23.186 | -2.313<br>(7.439) | -3.582<br>(10.689) | 194 |

Column 2 presents ANCOVA ITT effects using February - May 2020 as pre-treatment period and October 2020 - May 2021 as post-treatment period. Column 3 presents ANCOVA ATT effects on the same time periods using IV regressions with NHIF enrolment instrumented by treatment assignment. Column 5 and 6 present the simple aggregated ITT and ATT as proposed by Callaway and Sant'Anna (2021) using the cross-sectional sample and the doubly robust DiD estimator based on stabilized inverse probability weighting and ordinary least squares. All estimations include pair fixed effects and month fixed effects and standard errors are clustered at the pair level. \* $p < 0.1$ ; \*\* $p < 0.05$ ; \*\*\* $p < 0.01$ .

## Footnotes

1 The program also sought to stimulate savings for co-payment of next year’s health insurance premium through a digital savings component, the so-called “health wallet”, which runs on people’s mobile phone, using the M-TIBA platform. In our study area, the health wallet was not promoted to the participant households, and therefore not included in the evaluation.

2 NHIF enrollees are required to attend their preferred healthcare provider for out-patient care but are allowed to visit other NHIF-empaneled providers for in-patient or emergency care, or upon referral. Every quarter, enrollees get the opportunity to switch their preferred provider. This also applied to households enrolled through the digital insurance program.

3 Healthcare is free in public providers until they reach NHIF level 4, after which they are allowed to charge fees. Out of the 13 public healthcare providers in Khwisero sub-country, only one was in the process of upgrading to NHIF level 4 for empanelment in NHIF at the start of the study. Although it was only offering out-patient care at that stage, it had already started charging fees.

4 Women below 18 years old were excluded as target women from the study sampling frame because of ethical requirements, except pregnant girls aged 15 years and above who were considered emancipated minors.

5 We used the so-called “Euclidean distance methodology” for our matching process, which corresponds to the absolute difference between the standardized values of all of the covariates for a possible pair of matches.

6 Whereas the i-PUSH team assisted households in getting birth certificates for their children, it was not possible to assist adults in getting a national ID-card.

7 Outcome variables related to reproductive, maternal, neonatal and child health are reported elsewhere (Abajobir et al., 2023).

8 Taking the weekly average within months avoids dropping months for which one week is missing – essentially, we impute the monthly average for missing weeks.

9 The diaries data from December 2019 and January 2020 are excluded from the analysis because data collection was erratic during the Christmas period as many respondents were travelling. This also ensure high-quality data, allowing for on-the-job-learning during the early weeks of the diaries field work.

10 As a robustness check, we will also calculate impact estimates taking June as the start of the post-intervention period. This will provide a lower bound as program roll-out was only partial in the initial months.

11 The monetary poverty line is KShs 3,252 monthly per adult equivalent in rural areas (KNBS 2020). The average monthly income per capita our sample is 3600/4.9*4.2=3085KShs. The exchange rate on November 1^st^, 2019 (halfway the baseline survey) was 100 KES : 0,96 USD.

12 This excludes RMNCH visits for family planning, ANC, delivery, PNC, childhood immunization and childhood health check-ups/monitoring.

